# Metabolome-wide analysis suggests antidepressant exposure counteracts risk of depression in fatty-acid pathways

**DOI:** 10.64898/2026.11.20.26368585

**Authors:** Somying Tsai, Dennis Klose, Alex S.F. Kwong, Edward T. Bullmore, Golam M. Khandaker, Yuri Milaneschi, Brenda W.J.H. Penninx, Andrew M. McIntosh, Matthew Iveson, Heather C. Whalley, Xueyi Shen

## Abstract

Antidepressants are widely prescribed, yet their biological mechanisms remain incompletely understood. Large-scale metabolomics and linked electronic health records provide a unique opportunity to characterise metabolic profiles associated with medication exposure. We investigated metabolic signatures of antidepressant exposure using 251 nuclear magnetic resonance biomarkers in UK Biobank (N = 222,911), with external validation in NESDA and ALSPAC (N = 6,082) and longitudinal analyses (n = 5,820, mean follow-up = 4.2 years). We identified 152 metabolites associated with current antidepressant use (PFDR < 0.05), characterised by elevated triglycerides and very-low-density lipoproteins. Current users also showed higher omega-3 fatty acid levels and lower omega-6-to-omega-3 ratios, both associated with lower depression risk. Metabolic profile scores significantly distinguished antidepressant users in both external cohorts (OR = 1.07– 1.39, ROC AUC = 0.58–0.73). Longitudinal analyses identified nominal associations between antidepressant initiation and increases in nine lipid biomarkers (β = 0.144– 0.229). These findings suggest metabolic signatures associated with antidepressant use may not be solely explained by underlying depression and are characterised by both potentially beneficial fatty acid alterations and adverse lipid profiles. Validation across independent cohorts and consistency with longitudinal findings strengthen our evidence, supporting insights into antidepressant mechanism and its relevance to cardiometabolic health.

---

Major depressive disorder (MDD) is a debilitating psychiatric disorder with a high lifetime prevalence of approximately 10.8%^1^. The World Health Organisation has ranked depression among the largest contributors to global disability^2^. Antidepressants are the first-line pharmacological treatment for depression, with major classes including selective serotonin reuptake inhibitors (SSRIs), selective norepinephrine re-uptake inhibitors (SNRIs), and tricyclic antidepressants (TCAs)^3^. Antidepressant use has increased substantially in the global population^4,5^. For example, in England, 92.6 million antidepressant items were dispensed in 2024/25, representing an increase of nearly 50% over the past decade^6^.

Despite the increased use of antidepressants, low remission rates remain a significant challenge. Around 30-50% of patients with MDD are treatment-resistant^7–9^, and the biological mechanisms underlying the high variability in treatment response remain poorly understood. Improving clinical treatment therefore requires robust, replicable molecular evidence to help advance our understanding of the mode of action of antidepressants^10^. To date, few large population-scale molecular investigations of antidepressant use have been conducted. Previous findings have shown limited replication, and the causal effects of antidepressant use on molecular profiles remain underexplored.

Metabolic psychiatry is a rapidly emerging field that examines systemic metabolic abnormalities involved in the pathophysiology of mental health conditions^11^, and may provide novel mechanistic insights into antidepressants currently in clinical use. Up to 30% of patients with MDD show significant metabolic dysregulation, including abdominal obesity and dyslipidaemia^12^, and plasma metabolic biomarkers, such as fatty acids, have been causally implicated in risk of MDD^13–16^. Antidepressants, however, may further compound metabolic dysfunction. Weight gain and blood pressure disturbances are the most commonly observed side effects of antidepressant use that may lead to treatment discontinuation^17^. Specific antidepressants (e.g., duloxetine) are associated with cholesterol and glucose increase^18^. Despite growing evidence of treatment-related metabolic effects, existing studies have been limited to a narrow set of metabolites and/or broad clinical measures such as changes in weight and total cholesterol levels.

Recent advances in high-throughput metabolic profiling have enabled comprehensive metabolomic characterisation of antidepressant use. Beyond measuring total circulating lipid levels, nuclear magnetic resonance (NMR) spectroscopy enables detailed quantification of individual lipid components across different lipoprotein subclasses, such as triglyceride in high-density lipoproteins (HDL) and in low-density lipoproteins (LDL)^19^. NMR spectroscopy also quantifies circulating non-lipid metabolites, including glycoprotein acetyls and amino acids, providing insights into other metabolic pathways^19^. Notably, lipoprotein subclasses (e.g., HDL and LDL) differ in their relative proportions of lipid components (i.e., triglycerides, cholesteryl esters, phospholipids and free cholesterol), as well as in their biological functions and associations with cardiometabolic diseases^19–21^. Therefore, metabolome-wide analyses incorporating lipoprotein subclass-specific lipid measurements alongside non-lipid metabolites may provide a more comprehensive understanding of the metabolic signatures associated with antidepressant use.

Electronic health records (EHRs) provide rich, routinely collected clinical data that enable large-scale longitudinal measurement of medication dispensing^22^. Combined with advanced metabolomic profiling technologies, these data create opportunities to investigate metabolic changes associated with drug exposure. Metabolomic data linked to detailed clinical information remain scarce, with the UK Biobank (UKB)^19^, the Netherlands Study of Depression and Anxiety (NESDA)^23^, and the Avon Longitudinal Study of Parents and Children (ALSPAC)^24^ being the major extant cohorts that can provide large-scale NMR-based metabolomic data alongside EHR-derived information on the timing of antidepressant exposure. Importantly, plasma metabolites were measured at two timepoints in a subset of UKB participants^19^, supporting longitudinal assessment of metabolic changes and strengthening evidence for temporal associations^25^. Leveraging this unique opportunity, we used the UKB as a discovery cohort and sought to replicate our findings in NESDA and ALSPAC.

In our study, we utilised data from three large cohorts, the UKB, NESDA, and ALSPAC, to address the following aims: (1) to investigate metabolic signatures consistently associated with current antidepressant exposure by comparing current users with three control groups (never, past, and future users) in the UKB (N = 222,911, mean age = 56); (2) to interrogate the accuracy of the plasma metabolomic profiles in classifying individuals to antidepressant exposure status and antidepressant subclasses; (3) to validate the discovery results from UKB in two independent replication datasets, NESDA (N = 2,914, mean age = 41.9) and ALSPAC (N = 3,168, mean age = 24); and (4) to compare metabolic changes over time among participants who initiated antidepressants compared to sustained never users.

## Results

### Cross-sectional metabolome-wide association study (metWAS) in UK Biobank Case-control comparisons

This study integrated cross-sectional metWAS, replications using out-of-sample classification, and longitudinal analyses (Fig. 1). In the metWAS, 222,911 participants were classified into four antidepressant exposure groups using linked EHRs spanning up to 49.5 years: (i) current antidepressant users at the time of blood collection, (ii) never users with no record of antidepressant prescriptions, (iii) past users who had stopped antidepressants at least six months before baseline blood collection, and (iv) future users who initiated antidepressants after baseline blood collection (Table 1).

**Fig. 1.**
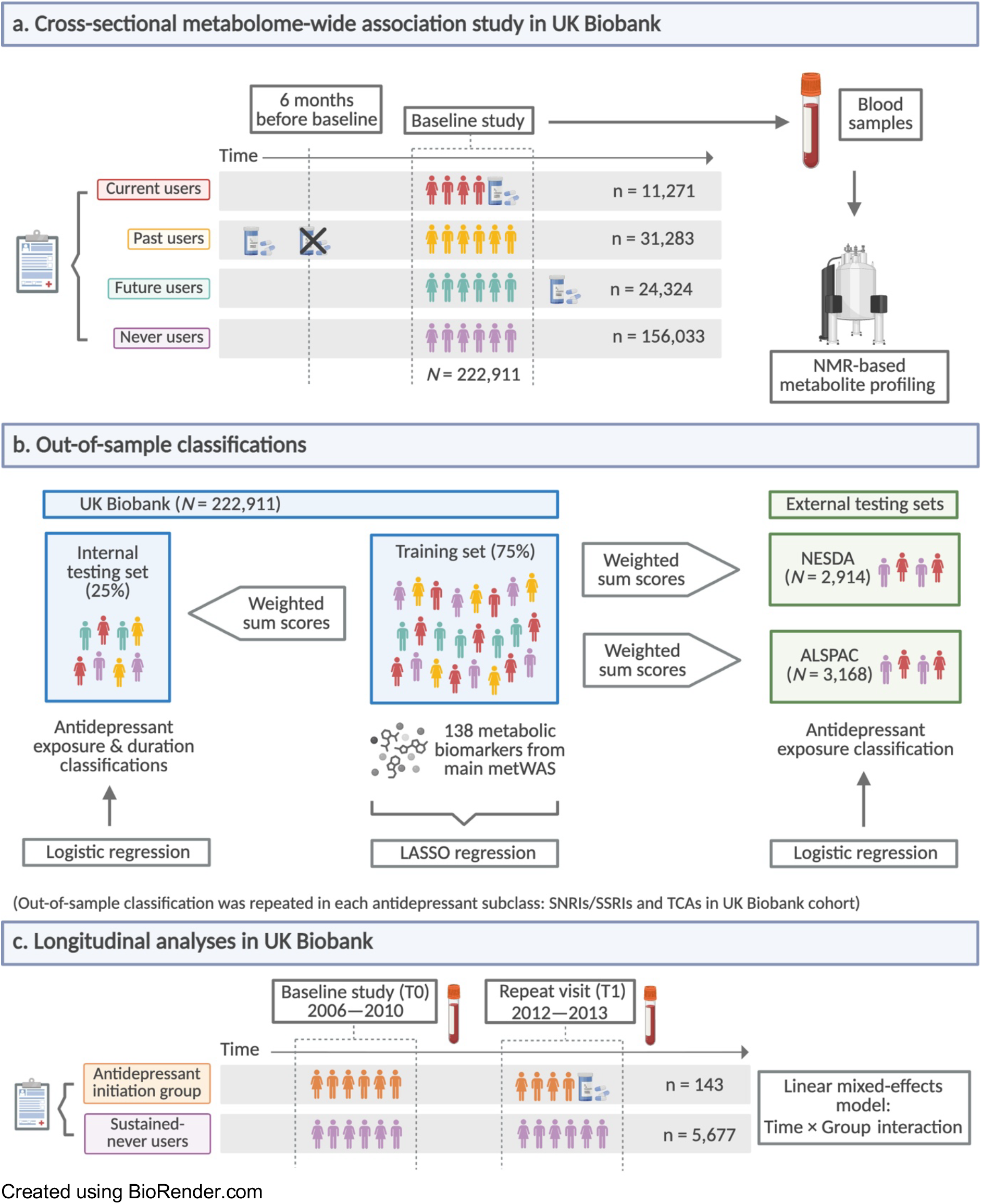
The main study design. (a) Cross-sectional metabolome-wide association analyses in UK Biobank using baseline data; (b) out-of-sample classifications using UKB as a discovery cohort and NESDA and ALSPAC as replication cohorts; and (c) longitudinal analysis in UK Biobank using metabolomic data collected during baseline and first repeat visit.

**Table 1.** Demographics of individuals in the cross-sectional metabolome-wide association analyses in UK Biobank using baseline data.

| **Demographic Characteristics** | **Current**<br>**n = 11,271** | **Never**<br>**n = 156,033** | **Past**<br>**n = 31,283** | **Future**<br>**n = 24,324** |
|---|---|---|---|---|
| **Age, Years, Mean (SD)** | 56.94 (7.78) | 56.49 (8.09) | 56.81 (7.88) | 56.21 (8.28) |
| **BMI, Mean (SD)** | 28.78 (5.59) | 27.32 (4.64) | 27.75 (4.97) | 27.83 (5.07) |
| **Female, %** | 68.95% | 50.31% | 65.49% | 60.94% |
| **SES, Mean (SD)** | -0.72 (3.32) | -1.48 (2.94) | -1.16 (3.09) | -0.98 (3.26) |
| **Never Smoked, %** | 48.03% | 56.77% | 50.52% | 51.76% |
| **Ethnicity (Mixed/Other/British), %** | 0.02%/0.63%/92.26% | 0.01%/0.73%/89.63% | 0.02%/0.73%/90.73% | 0.01%/0.76%/89.75% |
| **Had College/University Degree, %** | 22.92% | 35.08% | 25.64% | 27.60% |
| **Alcohol Intake Frequency (Daily or Almost Daily/Once or Twice a Week/One to Three Times a Month), %** | 16.39%/ 22.77%/ 12.86% | 20.62%/ 26.59%/ 10.88% | 17.78%/ 26.17%/ 12.22% | 18.06%/ 26.38%/ 12.07% |

BMI, body mass index; SES, socioeconomic status.

We conducted three case-control comparisons between current antidepressant users (cases) and three control groups: never users, past users, and future users. In the main models, we adjusted for technical covariates, age, sex, smoking status, socioeconomic status (SES), education, body mass index (BMI), ethnicity, alcohol drinking status, alcohol drinking frequency. Depression status (seen a general practitioner/psychiatrist for nerves, anxiety, depression)^26^ and depressive symptoms (Patient Health Questionaire-4) were also included as covariates to account for potential confounding by the underlying disease.

Of the 251, 152 metabolites were significant across all comparison models (Fig. S1 in Supplementary 1). The highest number of significant associations were found in the current/never comparison (189 significant associations, β = -0.164–0.179, *P_FDR_* = 1.83×10^-52^ to 0.0484), followed by the current/future comparison (177, β = -0.130– 0.150, *P_FDR_* = 8.29×10^-20^ to 0.049), then the current/past comparison (171, β = - 0.160–0.164, *Pfdr* = 1.35×10^-33^ to 0.0407). Full results are provided in Table S1—S3 in Supplementary 2. Effect sizes were significantly positively correlated across all three pairwise comparisons (r = 0.98–0.99, *P* < 2.2×10^-16^; Fig. S1). In the current/never comparison, 86% of the significant biomarkers measured lipids and lipoproteins (Fig. 2 and Fig. S2). In supplementary analyses, principal component analysis (PCA) was used to identify systems or dimensions of correlated metabolites associated with antidepressant exposure (further details are provided in Supplementary 1 and Table S4 in Supplementary 2).

**Fig. 2.**
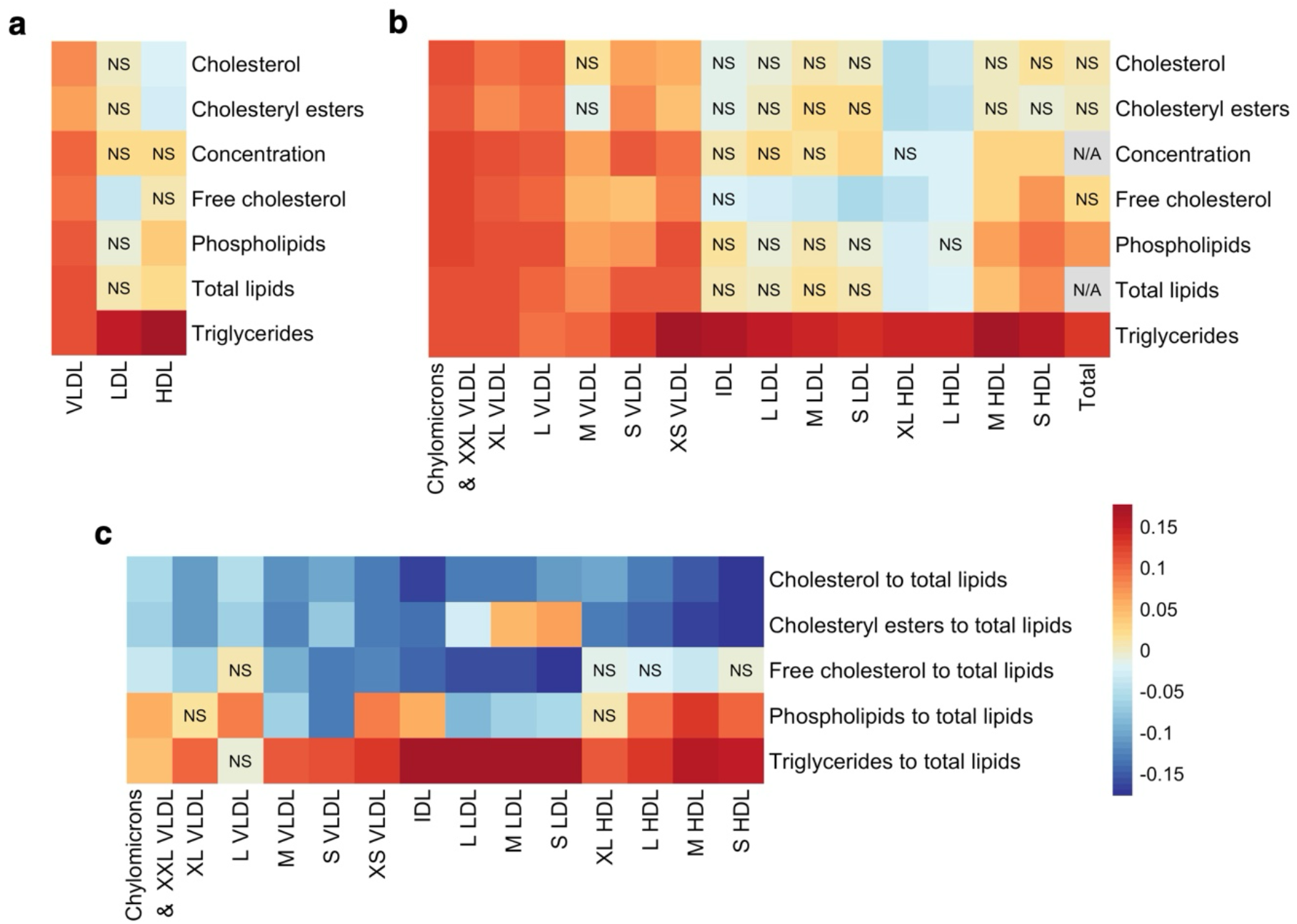
Heatmaps of effect sizes from the current/never comparison model. NS denotes non-significant (*P_FDR_* > .05) associations, and N/A denotes data not available. (a) Biomarkers measuring different lipid components of VLDL, LDL, and HDL. (b) 14 lipoprotein subclasses with their lipid component measured, along with total cholesterol, total cholesteryl esters, total free cholesterol, total phospholipids, and total triglycerides. (c) 14 lipoprotein biomarkers in the relative lipoprotein lipid concentration group. These heatmaps showed current users were associated with higher levels of triglycerides across all lipoprotein subclasses and higher levels of VLDL, compared to individuals who had never used antidepressants. VLDL, very low-density lipoproteins; IDL, intermediate-density lipoproteins; HDL, high-density lipoproteins; XXL, extremely large; XL, very large; L, large; M, medium; S, small; XS, very small

To evaluate whether the identified associations were driven by collider bias introduced by depression status or depressive symptoms, we conducted supplementary analyses using only age, sex, and the technical factors as covariates. The supplementary models (Table S5—S7 in Supplementary 2) showed largely consistent results, with 92% of metabolites significant in the main models remaining significant in the models with minimal adjustment for covariates (*P_FDR_* < .05).

### Sensitivity analyses

We conducted two sensitivity analyses to assess the robustness of our findings to depression status and cardiovascular health (Supplementary 1). After restricting the case-control comparisons to participants with depression, 98% of the significant biomarkers (149/152) remained significant (Table S8—S10 in Supplementary 2). After additional adjustment for cardiovascular risk factors in the main case-control comparisons (self-reported type 1 diabetes, type 2 diabetes, rheumatoid arthritis, atrial fibrillation, kidney failure, kidney nephropathy, glomerulonephritis, kidney nephropathy, glomerulonephritis, migraine, systemic lupus, and erectile dysfunction), all 152 significant biomarkers were retained (Table S11—S13 in Supplementary 2).

### Comparison to MDD study

We compared our current/never antidepressant use results with estimates of the causal effects of metabolites on MDD risk, as reported independently by Davyson et al.^13^ to explore whether antidepressant-associated metabolic differences may reflect the underlying associations implicated in MDD (Fig. 3a). Of the five biomarkers significantly implicated in MDD risk by these prior Mendelian randomisation (MR) analyses, three were also significantly associated with current antidepressant use: degree of unsaturation, omega-3 fatty acids, and the ratio of omega-6 fatty acids to omega-3 fatty acids (Fig. 3b). A higher degree of unsaturation was associated with lower MDD risk in MR analyses (β = -0.063, *P_FDR_* = 0.004), while current antidepressant users had a lower degree of unsaturation than never users (β = - 0.106, *P_FDR_* = 5.51×10^-22^). In contrast, higher omega-3 fatty acid levels were associated with lower MDD risk (β = -0.046, *P_FDR_* = 0.025), whereas current users had higher omega-3 fatty acid levels (β = 0.049, *P_FDR_* = 4.24×10^-5^). Similarly, a higher omega-6 to omega-3 fatty acid ratio was associated with increased MDD risk (β = 0.052, *P_FDR_* = 0.040), whereas current users had a lower ratio (β = -0.044, *P_FDR_* = 1.23×10^-4^). Overall, the directions of association were opposite between the MR and current/never analyses for omega-3 fatty acids and the omega-6 to omega-3 fatty acid ratio, whereas the direction was consistent for degree of unsaturation (Fig. 3).

**Fig. 3.**
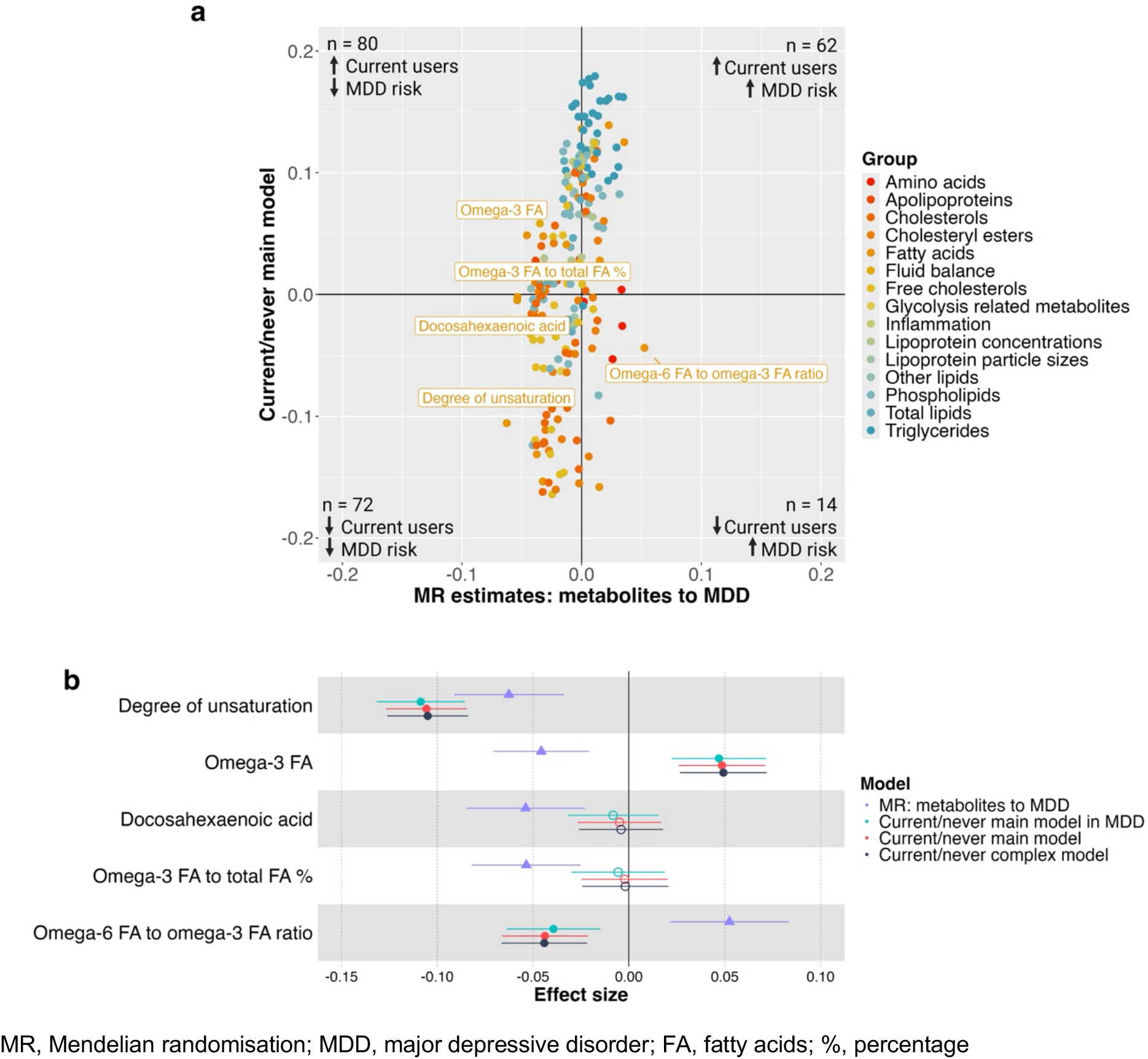
Comparison of effect sizes from current/never models with Mendelian randomisation (MR) estimates for MDD from Davyson et al^14^. (a) Scatter plot of main current/never model compared to MR estimates. Arrows indicate the direction of metabolite associations with current antidepressant users compared to never users and the direction of MR estimated causal effects of metabolite levels on MDD risk, respectively. The five significant biomarkers with potential causal effect on MDD were labelled. (b) Effect sizes from the main current/never model, the main current/never model restricted to MDD cases, and the complex current/never model adjusted for cardiovascular related covariates compared with inverse variance weighted MR estimates. Hollow dots denote non-significant association (*P_FDR_* > .05). Current users had higher omega-3 fatty acid levels and a lower omega-6 to omega-3 fatty acid ratio, both associated with lower MDD risk based on MR estimates.

We conducted supplementary analyses to further investigate the directional relationships between antidepressant-associated metabolic differences and the MR estimates of the causal effects of metabolites on MDD risk. We examined the distribution of metabolites showing the same or opposite directions of association between the MR estimates and current/never analyses across metabolite groups using chi-square goodness-of-fit and two-sided exact binomial tests (Table S1 in Supplementary 1). Overall, only the total lipid group showed a significantly higher proportion of opposite-direction associations in both tests, whereas the other lipid group reached significance only in the chi-square test.

### Metabolite set enrichment analysis (MSEA)

Lipoprotein subclass biomarkers were grouped according to their measured lipid components, while the original Nightingale classifications were retained for non-lipoprotein biomarkers, resulting in 16 metabolite groups. A full list of metabolites and groups can be found in Table S14 in Supplementary 2. Fourteen groups comprising 248 metabolites were included in the MSEA to investigate group-level enrichment (inflammation and fluid balance groups were excluded due to having fewer than three metabolites). The normalised enrichment scores (NES) were estimated using the effect sizes from the three case-control metWAS models.

Triglyceride group was positively enriched across all three comparisons (*P_FDR_* < .001; Fig. 4a), whereas cholesterol group was negatively enriched (*P_FDR_* < .05). Cholesteryl ester group was also negatively enriched in current users compared with never and past users (*P_FDR_* < .05), but not future users. These findings were consistent with the metWAS results (Fig. 2), indicating that current users generally showed higher triglyceride levels and a higher proportion of triglycerides relative to total lipids across lipoprotein subclasses. Amino acids were negatively enriched in current users compared with never and future users (*P_FDR_* < .05), but not past users.

**Fig. 4.**
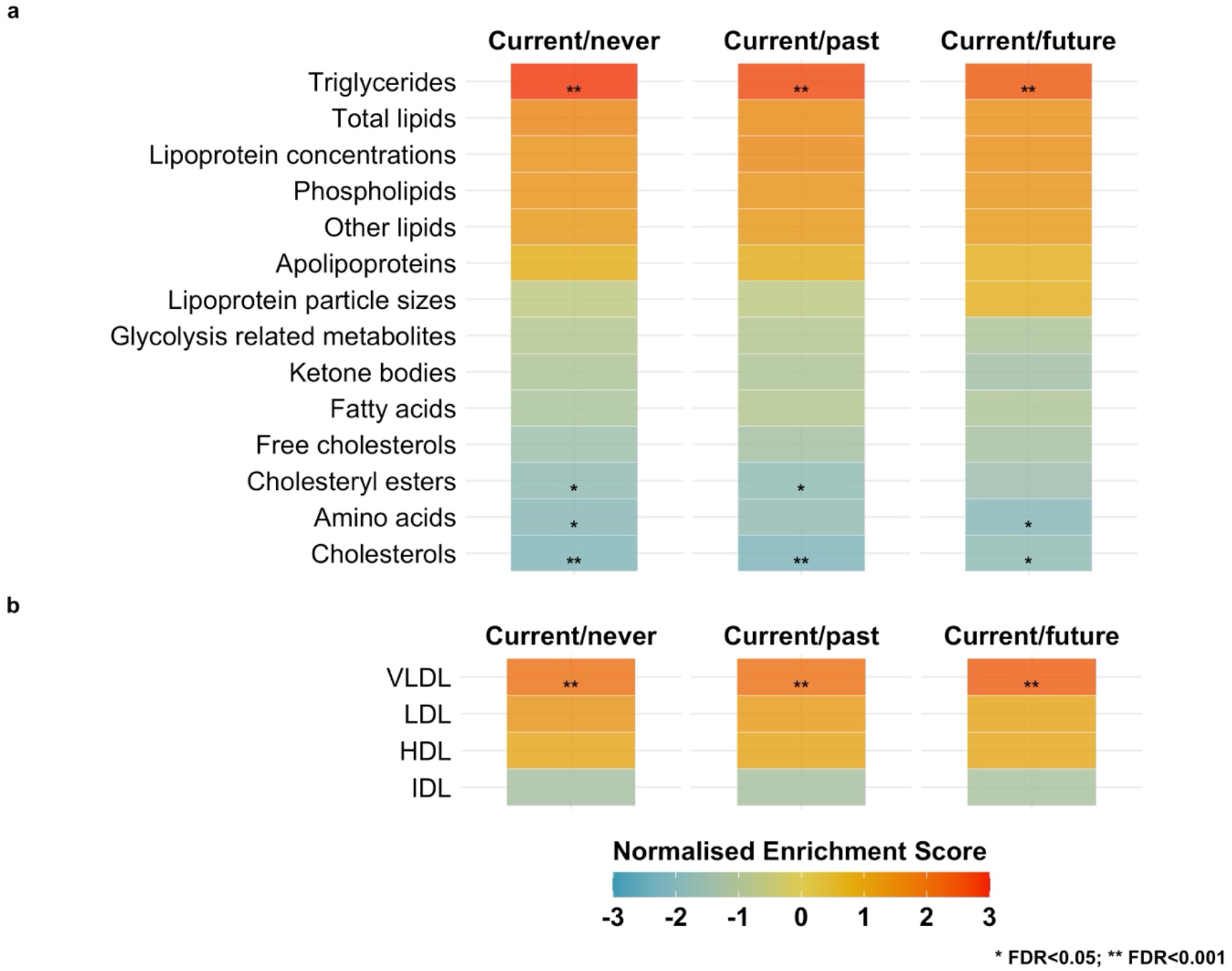
MSEA for (a) metabolite classes (N = 14) and (b) lipoprotein subclasses. Triglyceride group and VLDL group showed significant positive enrichment across all three exposure comparison groups. Cholesteryl esters, amino acid, and cholesterol groups showed significant negative enrichment in at least two comparisons. Metabolite Set Enrichment Analysis was used to estimate enrichment scores based on effect sizes from three main metWAS models. Multiple comparisons were accounted for using FDR. (a) We first analysed the overall metabolite groups (N = 14) and then (b) stratified the lipoprotein subclasses into biomarkers of very low-density lipoprotein (VLDL), low-density lipoprotein (LDL), intermediate-density lipoprotein (IDL), and high-density lipoprotein (HDL). *FDR < 0.05; **FDR < 0.001.

To examine overall enrichment patterns across lipoprotein subclasses, we additionally grouped lipoprotein biomarkers into VLDL, IDL, LDL, and HDL classes (Fig. 4b). For example, triglycerides in small HDL and triglycerides to total lipids in small HDL percentage were both assigned to the HDL group. VLDL biomarkers were positively enriched across all three case-control comparisons (*P_FDR_* < .001), consistent with the metWAS findings (Fig. 2), which identified higher levels of several biomarkers measuring VLDL and its lipid components. HDL and LDL biomarkers also showed positive enrichment, whereas IDL showed negative enrichment, although these associations did not reach statistical significance. Together, these findings suggest that the positive associations observed for individual triglyceride-and VLDL-related biomarkers in the metWAS reflect broader, consistent patterns of positive enrichment across the metabolite groups.

### Out-of-sample classifications using metabolic profile score

To validate the associations between metabolite abundance and current antidepressant exposure, we created metabolic profile scores in three independent samples: (1) a held-out sample in UKB sample (internal replication), (2) the Netherlands Study of Depression and Anxiety^27^ (NESDA; external replication), and (3) the Avon Longitudinal Study of Parents and Children^28–30^ (ALSPAC; external replication).

### Internal validation in UKB

Current and never users in UKB were split into training (75%) and testing sets (25%). In the training set, we repeated three case–control comparisons. A total of 138 metabolites showed significant differences between current users and all three control groups in the training set. These 138 metabolites were included in the LASSO (least absolute shrinkage and selection operator) regression, which identified 125 metabolite biomarkers with non-zero coefficients. A weighted sum score was generated using the LASSO weights. The score trained on the current and never users significantly classified all three comparison groups in the internal testing sample within UKB. The highest odds ratio (OR) was found for current/never group (OR = 1.38, ROC AUC = 0.59, PR AUC = 0.10, *P* < 2×10^-16^), followed by current/past (OR = 1.28, ROC AUC = 0.57, PR AUC = 0.11, *P* < 2×10^-16^, and current/future (OR = 1.27, ROC AUC = 0.57, PR AUC = 0.13, *P* < 2×10^-16^) (Fig. 5).

**Fig. 5.**
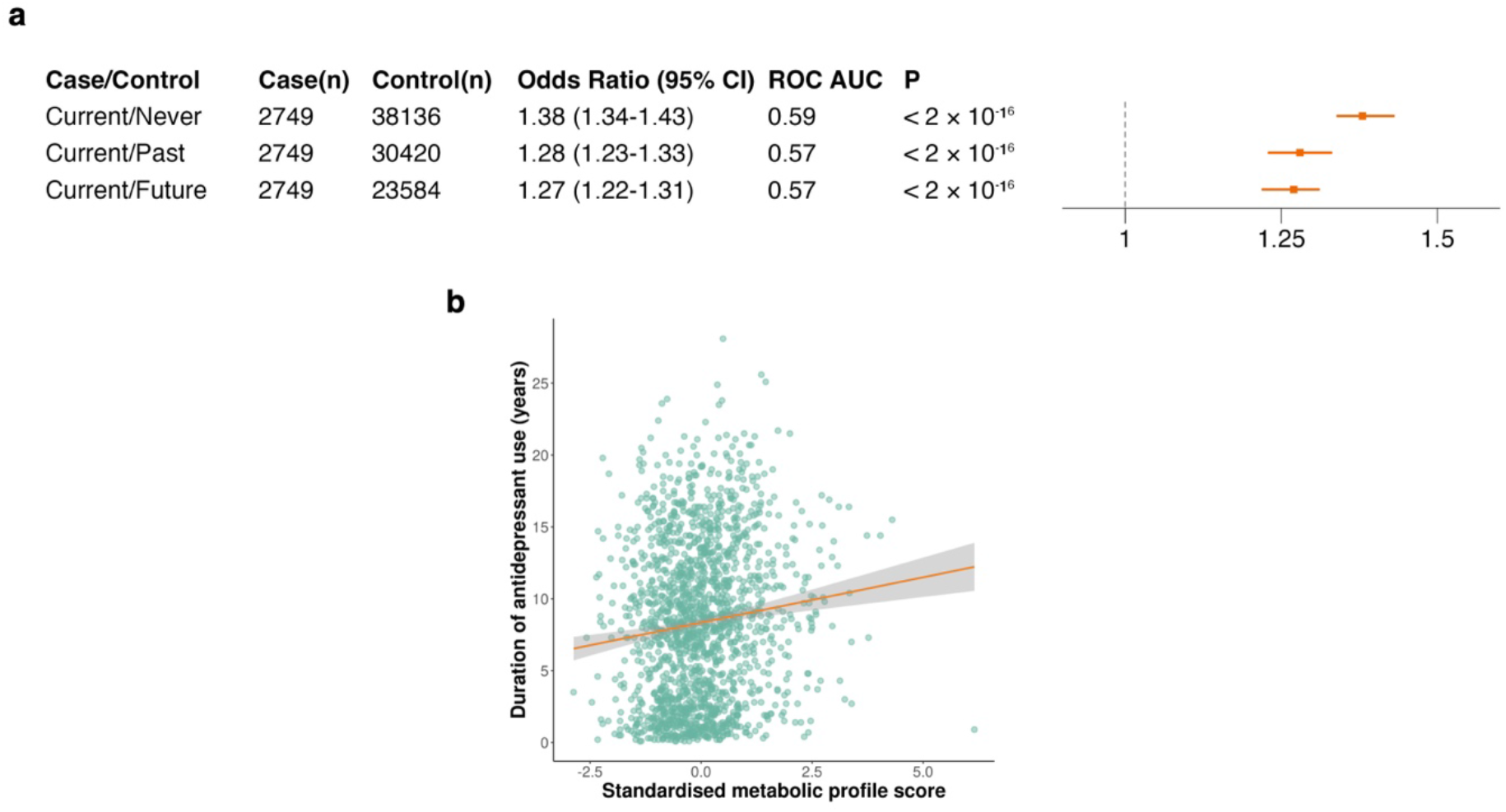
Out-of-sample classification using the metabolic profile score. (a) Internal validation of the score for classifying current antidepressant users and controls. (b) Scatter plot of standardised metabolic profile score and duration of current antidepressant use (years) among current users in testing set, with the fitted line from a linear regression model. The metabolic profile score distinguished current users from all control groups and higher score was associated with longer use of antidepressants.

### Duration of antidepressant exposure

Among current antidepressant users in the testing set (n = 1,608), we conducted a simple linear regression analysis to assess whether the metabolic profile scores were associated with the duration of antidepressant exposure (mean = 8.3 years). The score was significantly associated with duration of exposure (F(1, 1606) = 21.18, *P* = 4.51×10^-6^), with each 1-SD increase in the score associated with a 0.63-year increase in duration of exposure (95% CI: 0.49, 0.77; Fig. 5b). The score explained approximately 1.3% (R^2^ = 0.013) of the variance in exposure duration.

### External validation in different age cohorts

We leveraged two independent cohorts, NESDA (N = 2,914, mean age = 41.9) and ALSPAC (N = 3,168, mean age = 24), to externally validate the metabolic profile score in mid-life and young adult populations, respectively. We applied the LASSO coefficients estimated from the UKB training data to NESDA and ALSPAC to derive metabolic profile scores for individuals in each cohort.

#### Validation in mid-life adults

In NESDA, after adjusting for age, sex, and technical covariates, the metabolic profile score significantly discriminated current antidepressants users (n = 723) from non-current users (n = 2,191; OR = 1.18, ROC AUC = 0.63, *P* = 1.90×10^-4^; Fig. 6a). The association remained significant when the analysis was restricted to participants with current depression (OR = 1.14, ROC AUC = 0.58, *P* = 0.03) and after additional adjustment for smoking status, education, and BMI (OR = 1.13, ROC AUC = 0.73, *P* = 0.012). However, after both restricting the analysis to current depression cases and adjusting for these additional covariates, the association was no longer statistically significant (OR = 1.11, ROC AUC = 0.60, *P* = 0.116).

**Fig. 6.**
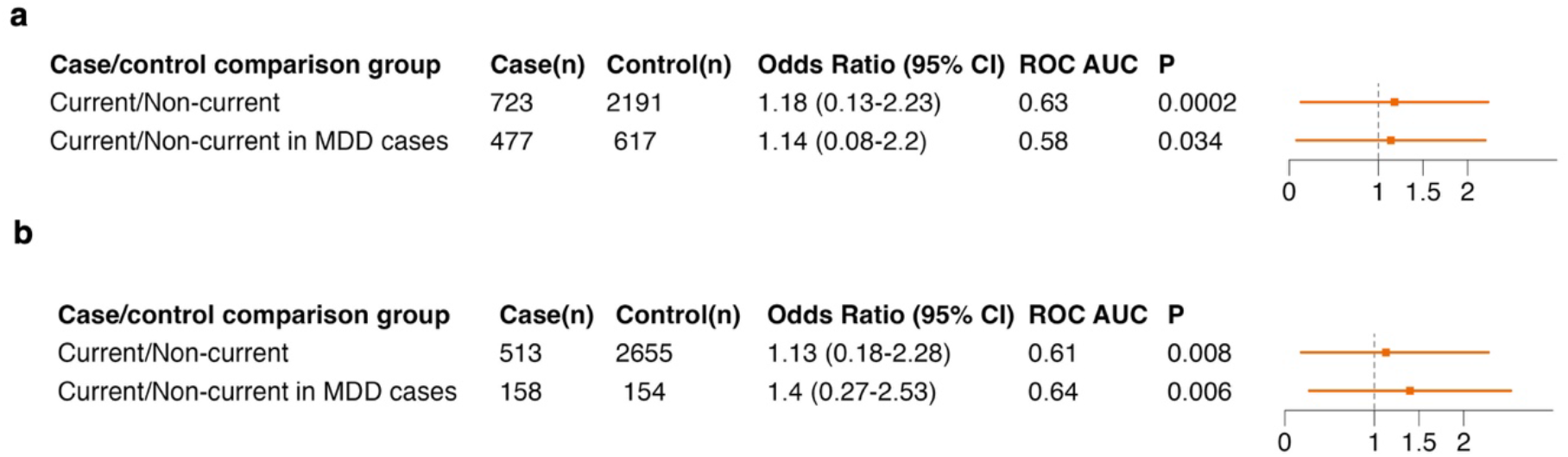
Out-of-sample classification using the metabolic profile score in NESDA and ALSPAC. (a) NESDA: Classification of current antidepressant users and non-current users in the full sample and among participants with MDD, adjusted for age, sex, and shipment. (b) ALSPAC: Classification of current antidepressant users and non-current users in the full sample and among participants with MDD, adjusted for sex.

#### Validation in young adults

In ALSPAC, we used two definitions of antidepressant exposure. In the primary definition, cases were current antidepressant users (n = 180) and controls were non-current users (n = 2,749), based on a self-reported questionnaire administered at age 24, when plasma metabolites were collected. The metabolic profile score did not significantly discriminate cases from controls (OR = 1.06, ROC AUC = 0.58, *P* = 0.413).

Given the small number of current users at age 24, we conducted additional analyses using a broader exposure definition incorporating a second self-reported item assessed at age 28. Cases (n = 513) were participants who reported either current antidepressant use at age 24 or prescribed medication for depression or anxiety within the previous five years at age 28. Controls (n = 2,655) were participants who responded “No” to both items. After adjusting for sex, the metabolic profile score significantly discriminated cases from controls (OR = 1.13, ROC AUC = 0.61, *P* = 0.008), including when restricted to participants with current depression (OR = 1.40, ROC AUC = 0.64, *P* = 0.006; Fig. 6b). After additional adjustment for smoking and BMI, the association was no longer statistically significant in the full sample (OR = 1.07, ROC AUC = 0.63, *p* = 0.161) but remained significant among participants with current depression (OR = 1.37, ROC AUC = 0.65, *P* = 0.014).

### Antidepressant subclasses: Case-control comparisons

The three case-control metWAS models (current/never, current/past, current/future) were repeated for each of the three antidepressant subclasses (Table S15—S23 in Supplementary 2). TCAs showed the greatest number of significant metabolites across comparisons of current users with the three control groups (153 metabolites), followed by SSRIs/SNRIs (138), while only four metabolites were significant across all three case-control comparisons in other types of antidepressants. Effect sizes from the current/never models for TCA and SSRI/SNRI use were strongly positively correlated (*r* = 0.91, *P* < 2.2×10^-16^; Fig. S3). In the current/never metWAS models, TCAs and SSRIs/SNRI users shared 153 significant metabolites, with 43 metabolites unique to TCA use and 20 uniques to SSRI/SNRI use. Effect sizes from the TCA and SSRI/SNRI current/never models are illustrated in Figs. S4—7.

We further compared our subclass-specific results with the MR estimates of metabolite effects on risk of MDD from Davyson et al.^14^ (Fig. 7). For omega-3 fatty acids and the omega-6 to omega-3 fatty acid ratio, current TCA and SSRI/SNRI users showed consistent results with those of non-class specific current/never and MR estimates. Subclass-specific differences were observed for docosahexaenoic acid (DHA) and the omega-3 fatty acids to total fatty acids percentage. Current TCA use was associated with higher levels of both biomarkers (β = 0.11, *P_FDR_* = 1.23×10^-9^ and β = 0.05, *P_FDR_* = 7.79×10^-3^, respectively), whereas higher levels were associated with lower MDD risk in the MR analyses (β = -0.05, *P_FDR_* = 3.85×10^-2^ and β = -0.05, *P_FDR_* = 2.51×10^-2^, respectively). In contrast, current SNRI/SSRI use was associated with lower levels of both biomarkers (β = -0.02, *P_FDR_* = 0.105 and β = -0.03, *P_FDR_* = 0.032, respectively), consistent with the non-subclass specific current/never comparison (β = -0.005, *P_FDR_* = 0.705 and β = -0.002, *P_FDR_* = 0.858) but opposite to the associations observed for TCA use.

**Fig. 7.**
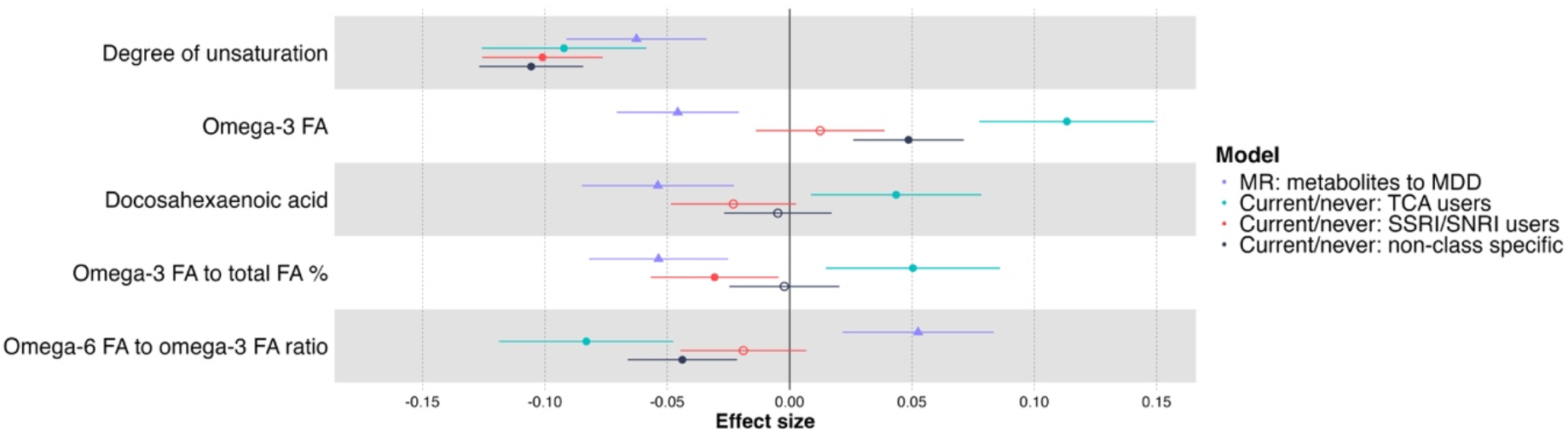
Forest plot comparing effect sizes from current/never models in SSRI/SNRI users and TCA users with MR estimates for MDD from Davyson et al^14^. Effect sizes from SSRI/SNRI users, TCA users, and non-class specific model were compared with inverse variance weighted MR estimates using metabolites as exposures and MDD as the outcome. Hollow dots denote non-significant association (*P_FDR_* > .05). Antidepressant subclass differences were observed for DHA levels and omega-3 fatty acid to total fatty acid percentage. Current TCA users had higher levels of both biomarkers, which were associated with lower MDD risk based on MR estimates, whereas current SSRI/SNRI users showed lower levels of both biomarkers. MR, Mendelian randomisation; MDD, major depressive disorder; FA, fatty acids; %, percentage

In supplementary analyses, we further compared the directions of association between MR estimates and antidepressant subclass-specific associations. We observed a higher proportion of significant opposite-direction associations across several metabolite groups, particularly for SSRIs/SNRIs, with some variation in directional patterns between SSRIs/SNRIs and TCAs (Table S2—S3 in Supplementary 1).

We also trained separate metabolic profile scores for SSRI/SNRI and TCA users using UKB data (Supplementary 1). In the internal UKB testing sample, both SSRI/SNRI- and TCA-trained scores significantly distinguished current SSRI/SNRI users and current TCA users from all three control groups, respectively. The SSRI/SNRI-trained score distinguished current TCA users from never users and the TCA-trained score distinguished current SSRI/SNRI users from never users, suggesting limited subclass specificity of the metabolic profiles.

### Longitudinal metabolomic analyses in UK Biobank

We leveraged longitudinal metabolomic data from UKB to further investigate metabolic changes following antidepressant initiation. A total of 5,820 participants had metabolomic data available at both baseline (T0) and follow-up (T1), with a mean interval of 4.33 years. Using linked EHRs, participants were classified into control group (never users at both timepoints) or antidepressant initiation group (non-current users at T0 who became current users at T1).

### Linear mixed-effect models

With the 152 metabolites associated with current antidepressant exposure in the cross-sectional analysis, we conducted linear mixed-effect models with time × group interaction adjusting for age, sex, BMI, month and time of blood collection. Nine metabolic biomarkers showed a nominally significant time × group interaction (β = 0.17–0.21, nominal *P* = 0.009 to 0.048), suggesting greater increases over time among the antidepressant initiation group compared with sustained-never users (Fig. 8a, Table S24 in Supplementary 2). However, none of the interactions survived FDR correction. The strongest interaction was observed for cholesteryl esters in very small VLDL (β = 0.21, *P* = 0.009), followed by cholesterol in very small VLDL (β = 0.20, *P* = 0.023), and the concentration of very small VLDL particles (β = 0.19, *P* = 0.020) (Fig. 8b). Of the nine significant biomarkers, eight measured lipids, and six measured lipid components and concentration of very small VLDL (Fig. 8a).

**Fig. 8.**
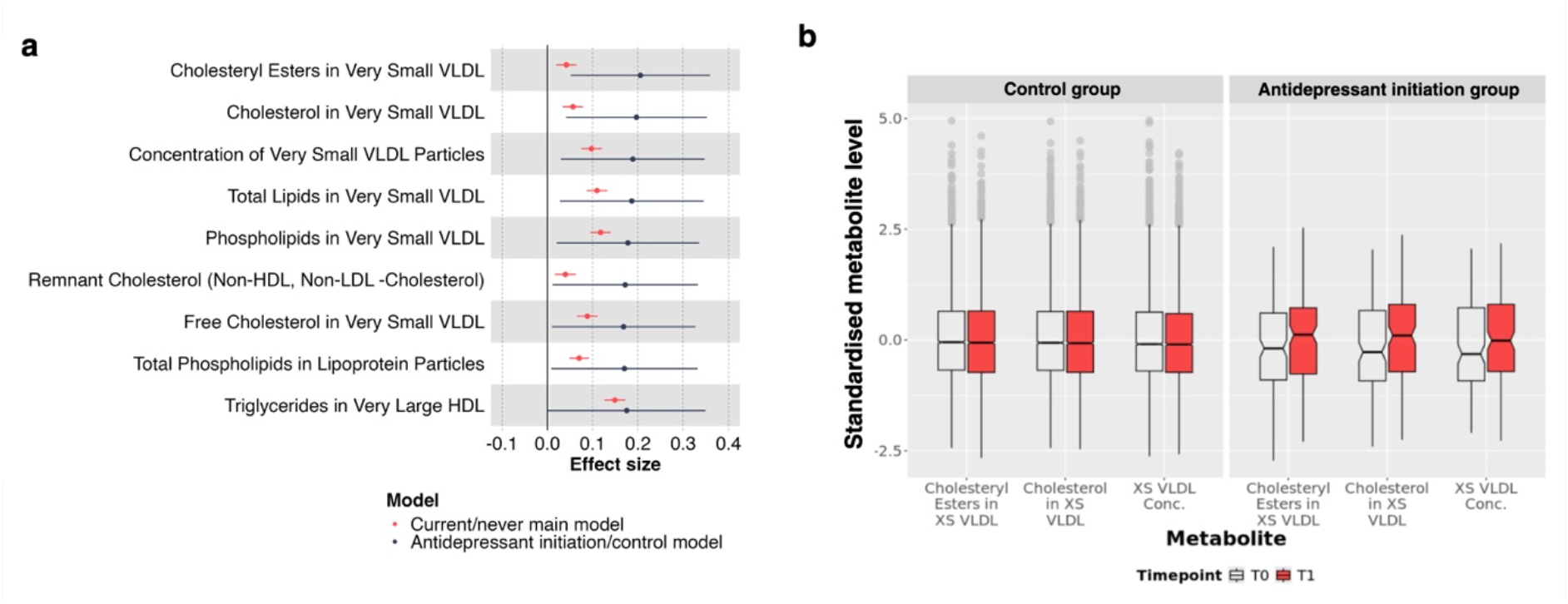
Longitudinal associations between antidepressant initiation and metabolite levels. (a) Forest plot comparing effect estimates for the nine nominally significant metabolites from the antidepressant initiation/control model with those from the current/never model. (b) Box plots of the three metabolites with the most significant time × group interactions. The box plots show the distributions of standardised metabolite levels at T0 and T1. Transparent boxes represent values at T0, and red boxes represent values at T1. The horizontal line within each box indicates the median, the notch represents the 95% confidence interval (CI) of the median, and grey dots denote outliers XS, very small; VLDL, very low-density lipoproteins; Conc., concentration

In addition, we applied the parametric g-formula to estimate the potential causal effects of antidepressant exposure on metabolite levels using the longitudinal data (further details can be found in Supplementary 1). We found nine nominally significant biomarkers that were directionally consistent with cross-sectional metWAS (Table S25 in Supplementary 2). Seven of which measured lipids and concentration of chylomicrons and extremely large VLDL.

Linear mixed-effect models and parametric g-formula analyses showed consistent directions of association, although the individual biomarkers reaching nominal significance differed between methods. Both analyses identified associations involving VLDL.

## Discussion

To our knowledge, this is the largest metabolome-wide association study of antidepressant exposure to date, examining 251 plasma metabolites with detailed characterisation of lipoprotein subclasses, lipid composition, and other metabolic biomarkers. We identified 152 metabolites uniquely associated with current antidepressant use across comparisons with three control groups, after accounting for depression status, depressive symptom severity, and lifestyle factors. Notably, 92% of metabolites significant in the main models remaining significant in minimally adjusted models including only age, sex, and technical covariates. Moreover, 98% remained significant in analyses restricted to individuals with depression, while all 152 remained significant after additional adjustment for cardiovascular risk factors. Together, these findings suggest that the associations were largely robust across different model specifications. Comparisons with a previous metabolite-MDD study^13^ showed both similarities and differences in metabolic associations, suggesting that the observed signatures may not be solely explained by underlying depression, although some may reflect biological pathways relevant to depression risk.

Our findings were further supported by out-of-sample classification in two independent cohorts spanning different age groups: NESDA (mean age = 41.9 years) and ALSPAC (mean age = 24 years). In both cohorts, the metabolic profile scores discriminated current from non-current users (ROC AUC 0.58–0.73). Higher scores were also significantly associated with longer duration of current antidepressant use. Furthermore, longitudinal analyses in UKB showed greater increases in nine lipid and lipoprotein biomarkers among antidepressant initiators compared with sustained-never users. All nine were directionally consistent with the cross-sectional metWAS findings and supplementary parametric g-formula analyses. Although these associations did not survive FDR correction, they provided potential temporal evidence for antidepressant-associated metabolic changes over time. Collectively, the external validation, associations with exposure duration, and longitudinal findings provide complementary evidence supporting the robustness of the observed metabolic signatures.

An important consideration is whether the observed metabolic signatures reflect antidepressant exposure rather than MDD itself. A previous study identified several fatty acid biomarkers with evidence of potential causal effects on MDD^13^. Notably, current users had higher omega-3 fatty acid levels and a lower omega-6 to omega-3 fatty acid ratio, both in directions associated with lower MDD risk based on MR estimates. These findings may be relevant to brain and cardiovascular health, as omega-3 fatty acids have been associated with neuronal preservation and protection against neurodegeneration, as well as lower cardiovascular mortality and major adverse cardiovascular events^31–34^. Conversely, a high omega-6 to omega-3 fatty acid ratio has been associated with cardiovascular disease, cancer, and inflammatory diseases^35^. However, similarities between antidepressant-associated metabolic signatures and MR estimates for MDD were also observed. Current users showed a lower degree of unsaturation, while MR estimates suggested that lower levels were associated with higher MDD risk. Future research is needed to determine whether these similarities reflect effects of antidepressant use or shared biological pathways underlying depression.

Subclass analyses further suggest that these fatty acid signatures may differ between antidepressant classes. Despite broadly similar metabolic profiles between SSRI/SNRI and TCA users, class-specific differences were observed for docosahexaenoic acid (DHA) and omega-3 fatty acids to total fatty acids percentage. Current TCA use was associated with higher levels of both biomarkers, which were associated with lower MDD risk based on MR estimates, while current SNRI/SSRI use was associated with lower levels of both. These differences perhaps partly reflect drug-specific effects on appetite and eating behaviours. Previous studies have reported differences in appetite and weight gain across antidepressants^36,37^. For example, TCAs such as amitriptyline and nortriptyline, have been linked to weight gain, increased appetite and craving for carbohydrates^38,39^, whereas venlafaxine (an SSRI) does not have a significant effect on weight^36^. Findings for SSRIs are more heterogenous^37^. These effects may contribute to the observed differences in metabolic profiles between subclasses, although further investigation is required.

Previous studies have consistently associated antidepressant use with higher plasma triglycerides^40–42^, a lipid marker associated with increased cardiovascular risk^43,44^. We extend these findings by showing that current antidepressant users had higher triglycerides concentrations across VLDL, LDL, and HDL subclasses. Moreover, higher concentration across all VLDL lipid components further suggest that these associations extend beyond triglycerides to broader alterations in lipid composition. This pattern was further supported by longitudinal analyses, in which antidepressant initiators were associated with greater increases in VLDL lipids.

While HDL cholesterol is inversely associated with cardiovascular risk^45–47^, higher triglyceride content within HDL has been associated with coronary artery disease, myocardial infarction, and ischemic stroke^48,49^, and may impair HDL structure and its anti-inflammatory and antioxidant properties^50,51^. We also observed higher levels of glycoprotein acetyls, a marker of chronic low-grade inflammation^52^, and apolipoprotein B, which is implicated in the development of atherosclerosis and plaque formation^53–55^. Conversely, apolipoprotein A1, which has established anti-inflammation properties^56,57^, was also higher among current users. Together, our findings suggest wider disturbance in lipoprotein lipid composition, particularly involving triglyceride and VLDL, along with alterations in biomarkers relevant to inflammation and cardiovascular health.

There is growing evidence linking both MDD and antidepressant use to metabolic syndrome. A recent review estimated that immune-metabolic dysregulation is present in around 20-30% of individuals with MDD^12^. The metabolic signatures associated with antidepressant use in our study also showed similarities to comparable results previously reported in metabolic syndrome. Specifically, elevated circulating triglycerides and glucose are established features of metabolic syndrome^58^. This overlap is notable given that weight gain, increased BMI and glucose levels, and atherogenic dyslipidaemia are recognised adverse effects of several antidepressants^40,59,60^ and are closely linked to metabolic syndrome^61–64^. Previous research has linked antidepressant use to metabolic syndrome even after accounting for MDD and depressive symptom severity^65–67^, with some studies reporting stronger associations among TCA users^66,67^. Consistent with this, we observed more extensive metabolic associations among TCA users than among SSRI/SNRI users.

Antidepressant utilisation among young people has increased substantially^6,68–70^. For example, in England, 7.9 million antidepressant items were dispensed to individuals aged 15-29 in 2024/25, representing a nearly 68% increase over the past decade^6^. Meta-analyses of antidepressant use among children and young adults under 25 have also raised concerns about an increased risk of suicidal ideation and behaviours^71,72^. However, the metabolic effects associated with antidepressant use in this population remain poorly characterised. Here, we showed antidepressant-associated metabolic profiles in ALSPAC participants at age 24 (OR=1.13—1.39, ROC AUC = 0.61—0.65, *P* < 0.05). These findings suggest that some of the metabolic signatures associated with antidepressant use in older adults may also present in younger populations, consistent with previous studies linking SSRI use in young adults to increase BMI and a small increased risk of type 2 diabetes^73,74^. However, the ALSPAC validation sample was modest, especially when the analyses were restricted to individuals with MDD. Furthermore, while metabolic profiles were measured at age 24, the antidepressant exposure definition in ALSPAC incorporated retrospective self-reported data collected at age 28. Therefore, these findings warrant further investigation in larger young-adult cohorts.

This study leveraged large-scale data, out-of-sample validations, and longitudinal data. However, several limitations should be considered. The imbalance between case and control sample sizes may have affected the classification models and contributed to their modest discriminative performance^75^. However, the precision-recall area under the curve (PR AUC), which is particularly informative for imbalanced dataset^76^, exceed that of a naive classifier across all models. Another limitation is the potential influence of health-related and lifestyle confounders on the observed associations. To mitigate this, we included an extensive set of covariates in our main metWAS models and conducted sensitivity analyses accounting for MDD, lifestyle factors, and cardiovascular risk factors. In addition, reliance on a single baseline metabolomic measurement may not fully capture long-term metabolic patterns. We therefore utilised repeat assessment data to examine longitudinal associations between antidepressant exposure and metabolic changes, providing complementary evidence to the single cross-sectional findings. Finally, the predominantly European ancestry of the UKB population may limit the generalisability of our findings to other ethnic groups.

In summary, this study characterised metabolomic signatures associated with current antidepressant use, identifying higher triglycerides across lipoprotein subclasses and alterations in VLDL lipid components, with stronger associations observed among long-term users. These findings were supported by external validation in two independent cohorts and longitudinal analyses. Comparisons with previous MDD study revealed both similar and different metabolic associations, indicating both shared and distinct biological pathways. In particular, current users had higher omega-3 fatty acid levels and lower omega-6-to-omega-3 ratios, both associated with lower MDD risk, suggesting that antidepressant exposure may shift fatty acid profiles in directions that counteract metabolic risk factors for MDD. Our findings warrant further investigation into the potential value of monitoring metabolic profiles among patients prescribed antidepressants and contribute towards a better understanding of the impact of antidepressant use on metabolic health.

## Methods

### Study population

UKB is a large cohort study comprising over 500,000 participants recruited across the United Kingdom, aged 40-69 years^19^. Baseline assessments were conducted between 2006 and 2010. A subset of participants underwent a repeat assessment between 2012 and 2013^19^. We included 222,911 participants with baseline metabolomic profiling data and linked EHRs for the cross-sectional metWAS analyses. We then included 5,820 participants with both metabolomic data available at baseline and follow-up for longitudinal analyses. Ethical approval for UKB was obtained from the North West NHS Research Ethics Committee (11/NW/0382). Access to the data for this study was granted under application #4844.

### Metabolic biomarkers

Metabolomic profiling of plasma samples from UKB was performed by Nightingale Health Plc. using the high-throughput NMR spectroscopy platform. Detailed procedures of the Nightingale NMR pipeline have been described elsewhere^19^. A total of 251 metabolic biomarkers were derived from non-fasting ethylenediaminetetraacetic acid (EDTA) plasma samples, including absolute concentrations and ratios of circulating metabolites, lipids, and lipoprotein sub-fractions^19^. Nightingale grouped these biomarkers by types into 18 categories: amino acids, apolipoproteins, cholesterol, cholesteryl esters, fatty acids, fluid balance, free cholesterol, glycolysis related metabolites, inflammation, ketone bodies, lipoprotein particle concentrations, lipoprotein particle sizes, lipoprotein subclasses, other lipids, phospholipids, relative lipoprotein lipid concentrations, total lipids, and triglycerides^19^. A full list of metabolic biomarkers, groups, and formulae for deriving the composite biomarkers and ratios is provided in Table S26 in Supplementary 2.

In this study, we regrouped the metabolic biomarkers to facilitate comparisons of lipid components across lipoproteins. Biomarkers of lipoproteins were grouped according to their measured lipid components, while the original Nightingale classifications were retained for non-lipoprotein biomarkers, resulting in 16 metabolite groups. For example, cholesterol in large HDL was assigned to the cholesterol group rather than the lipoprotein subclass group, while cholesteryl ester in IDL was assigned to the cholesteryl ester group. A full list of the derived metabolite groups is provided in Table S14 in Supplementary 2.

### Assessment of antidepressant exposure from electronic health records (EHRs)

Antidepressant exposure was defined using linked prescription and registration records from general practitioners (GPs) for 222,911 participants with available metabolomic profiling data. Prescription records included drug name, classification, quantity, and prescription date. Antidepressant records were extracted using drug names and Read v2/British National Formulary (BNF) codes, following the procedure established by Fabbri et al. (2021)^77^.

Following the pipeline created by Davyson et al.^78^, antidepressant treatment episodes were defined as at least two consecutive prescriptions with gaps of less than six months. Prescriptions occurring more than six months after the previous record were considered the start of a new episode, and single prescriptions of antidepressant were excluded. Participants with available metabolomic data were categorised based on whether they had an antidepressant episode during, before, or after the baseline assessment:

1. Current users (N = 11,271): had an antidepressant episode during the date of baseline assessment;
2. Past users (N = 31,283): had no current antidepressant episode, but had at least one past episode at least six months before the baseline assessment;
3. Future users (N = 24,324): had no current or past antidepressant episode, but had at least one future episode started after the baseline assessment;
4. Never used (N = 156,033): had a GP registration record but had no record of antidepressant use.

### Metabolomic data imputation

Missing metabolomic data were imputed using the ‘missRanger’ package (version 2.6.1)^79^ in R, which combines random forest prediction with predictive mean matching. Random forests have demonstrated superior performance compared to other machine learning models^80^ and have been applied to other omics data^57^. After removing individuals with missing age and sex data and those with more than 60% missing metabolites, 488,083 individuals were included in the imputation. We specified 50 trees per forest to balance computational efficiency and predictive performance. Predictive mean matching with three nearest neighbours was implemented to preserve the original data distribution and reduce imputation bias by replacing predicted values with observed values from similar cases^82^.

### Cross-sectional metabolome-wide association study (metWAS) in UK Biobank Case-control comparisons

We conducted three case-control comparisons between current antidepressant users (cases) and three control groups: past users, future users, and never users. The association between each metabolite and antidepressant exposure was assessed using general linear regression. Metabolites that were consistently significant across all three comparisons (current/never, current/past, and current/future) were considered uniquely associated with current users. Metabolites were standardised using z-score transformation to allow direct comparison of effect sizes across models. Therefore, all reported beta coefficients represent the standardised mean difference (Cohen’s d) between cases and controls. P-values were corrected for multiple testing using the false discovery rate (FDR) across all metabolic biomarkers within each case-control comparison (*P_FDR_*).

### Covariates

In the main models, we included age, sex, assessment centre, NMR spectrometer, SES, smoking status, ethnicity, educational attainment, BMI, alcohol drinker status, and alcohol intake frequency as covariates. Depression diagnosis^26^ and depressive symptoms severity (PHQ-4) were also included to account for potential confounding by MDD. See supplementary 1 for details about covariates.

### Sensitivity analyses

Two additional sensitivity analyses were conducted:

1. Restricting the main metWAS models to participants with depression only, to account for potential confounding between healthy and affected individuals.
2. Further adjusting for risk factors included in calculating cardiovascular risk^83^ including type 1 and type 2 diabetes, rheumatoid arthritis, kidney failure, kidney nephropathy, glomerulonephritis, migraine, systemic lupus, and erectile dysfunction, to ensure that metabolite associations were not fully driven by differences in cardiovascular risk. Further details about the covariates are provided in the Supplementary 1.

### Comparison to MDD study

To further compare the metabolic associations observed in our study to those reported for MDD^13^, we examined the distribution of effect directions across metabolite groups using chi-square goodness-of-fit and two-sided exact binomial tests.

### Metabolite set enrichment analysis (MSEA)

To investigate whether specific metabolite groups were enriched, MSEA was performed using ‘fgsea’ package (version 3.23)^84,85^ to estimate enrichment scores based on the effect sizes from the main metWAS models. To examine overall enrichment patterns across lipoprotein subclasses, we additionally grouped lipoprotein biomarkers according to four major lipoprotein classes: VLDL, IDL, LDL, and HDL classes. For example, triglyceride in very small VLDL was assigned to VLDL for this MSEA. A full list of metabolites in each group is provided in Table S14 in Supplementary 2.

In MSEA, effect estimates for each metabolite set were summarised using a normalised enrichment score (NES)^86^. The NES reflects the degree to which a set of metabolites is overrepresented at the top (positive effect sizes) or bottom (negative effect sizes) of the ranked list of metabolites. The NES is normalised to account for the differences in set size and statistical significance is assessed by controlling the false discovery rate.

### Out-of-sample classifications using metabolic profile score Internal validation in UKB

Following the association analyses, out-of-sample classifications were conducted to validate the classification potential of metabolic biomarkers in case-control comparisons using independent datasets. In the UKB baseline data, participants with current/never antidepressant status were split into training (75%) and testing (25%) sets. The ’caret’ R package (version 7.0-1)^87^ was implemented to ensure balanced representation of cases and controls. The three main metWAS models (current/never, current/past, current/future) were repeated in the training set, and only the metabolites consistently significant across all three case-control comparisons in the training set were included in the classification analyses. The metabolite values in both training and testing sets were standardised and residualised against covariates before further analyses. Covariates included age, sex, assessment centre, NMR spectrometer, SES, smoking status, ethnicity, educational attainment, BMI, alcohol drinker status, and alcohol intake frequency.

Penalised logistic regression using the least absolute shrinkage and selection operator (LASSO) was applied to the metabolite residuals, with current/never status as the binary outcome. Model hyperparameters were optimised via repeated threefold cross-validation (five repeats). To address class imbalance, random over-sampling examples (ROSE)^88^ was applied. LASSO coefficients reflected the relative importance of each metabolite in classifying the outcome^89^. Non-zero coefficients from the training set were multiplied by corresponding residualised metabolite values in the testing set to generate a weighted composite metabolic score per participant. Then, logistic regression was applied to use the score to classify current/never, current/past, and current/future status in the testing set. Areas under the curve (AUCs) and ORs were reported for classification performance^90^. The score was standardised using z-score transformation, therefore the reported OR represented the change in odds associated with a one standard-deviation (SD) increase in the score. Additionally, among current antidepressant users in the testing set, a linear regression analysis was conducted to assess whether the metabolic profile score was associated with the duration of antidepressant exposure.

### External validation in different age cohorts

Two external cohorts were included as independent test sets to evaluate the reproducibility and generalisability of our trained model: NESDA^27^ (mean age = 41.9) and ALSPAC^29^ (mean age = 24). In each cohort, the non-zero coefficients from the UKB LASSO training were multiplied by corresponding metabolite values to generate a weighted composite metabolic score per participant. The score was standardised using z-score transformation. Further details about the external cohorts are provided in the Supplementary 1.

In NESDA, participants were classified as current antidepressant users (n = 723; case group) or non-current users (n = 2,191; control group) based on drug container inspection of medications used in the past month, with medications classified according to the Anatomical Therapeutic Chemical classification system. The metabolic profile score was used to classified current and non-current users, adjusting for age, sex, and shipping batch.

In ALSPAC, participants were initially classified using a self-reported data on current antidepressant use at the time of blood collection during a research clinic visit age 24. Participants were categorised as current users of antidepressant at age 24 (n = 180; case group) or non-current users (n = 2749; control group). To increase statistical power, we conducted additional models incorporating data from a second self-reported questionnaire administered at age 28, which asked participants whether they had been prescribed medication for depression or anxiety within the past five years. In these additional models, the case group (n = 513) was defined as a “Yes” response to either: (1) antidepressant use at age 24, and/or (2) prescription of medication for depression or anxiety within the past five years, assessed at age 28. The control group (n = 2,655) was defined by negative responses to both self-reported items. The metabolic profile score was used to classified current and non-current users, controlling for sex. Following adjustment for basic covariates, we conducted sensitivity analyses in both external cohorts by further adjusting for smoking, education, and BMI.

### Antidepressant subclasses: Case-control comparisons and out-of-sample validation

In the UKB cohort, case-control metWAS and out-of-sample validation analyses were repeated for each antidepressant subclass to identify shared and class-specific metabolic signatures. Antidepressants were categorised into three groups: 1) SSRIs/SNRIs, 2) TCAs, and 3) other antidepressants. Categorisation was based primarily on BNF codes^77^, with drug names used as a supplementary resource when BNF codes were unavailable. Prescription records and treatment episodes were derived separately for each subclass:

- SSRIs/SNRIs: current (n= 6,729), past (19,072), future (13,254), never (183,223)
- TCAs: current (n= 4,221), past (20,815), future (18,387), never (178,855)
- Other antidepressants: current (n= 459), past (3,890), future (3,916), never (214,013)

We also compared the metabolic associations observed in SSRIs/SNRIs and TCAs to those reported for MDD^13^. We examined the distribution of effect directions across metabolite groups using chi-square goodness-of-fit and two-sided exact binomial tests.

### Longitudinal metabolomic analyses in UK Biobank

The final stage of this study incorporated data from the UKB baseline and repeat assessments, including both NMR-based metabolomic profiles and linked EHRs, to examine longitudinal associations between antidepressant exposure and metabolic patterns. A total of 19,877 participants had linked EHRs and metabolomic data available at both baseline and repeat assessment. Missing metabolomic data at the repeat assessment were imputed using the same approach described in the cross-sectional metWAS.

Antidepressant exposure at the repeat assessment was defined using the same procedure as in the cross-sectional metWAS^78^. Participants were categorised as current (n = 343), past (n = 58), future (n = 0), or never (n = 5,677) users based on antidepressant treatment episodes at the repeat visit.

Based on antidepressant status at both baseline and repeat visits, participants were further classified into two groups:

1. Control group (n = 5,677): participants who had a GP registration record but no record of antidepressant use in both baseline and repeat visits.
2. Antidepressant initiation group (n = 143): participants who were not current antidepressants users at baseline but had a current antidepressant episode at the time of the repeat assessment.

### Linear mixed-effects model

We applied linear mixed-effects models to investigate whether changes in metabolite levels over time differed between controls and antidepressant initiators. Only the 152 significant metabolites identified across the three main metWAS models were included. Random intercepts were included to account for repeated measurements within individuals, with fixed effects for time (T0 = baseline, T1 = repeat assessment), group (controls, antidepressant initiators), and their interaction (time × group).

Covariates included age, sex, BMI, spectrometer, month of blood collection, and time of blood collection. Time of blood collection was categorised as morning (05:00 ≤ 12:00), afternoon (12:00 ≤ 17:00), evening (17:00 ≤ 21:00), or night (21:00 ≤ 05:00). Analysis was conducted using ‘lmerTest’ R package (version 3.2)^91^. All metabolite values at T0 and T1 were standardised together by z-score transformation. The time × group interaction coefficient therefore represented the difference in the standardised change in metabolite levels over time between control group and initiator group.

## Data Availability

The data used in this study is available from UK Biobank with restrictions applied. Data were used under license and thus not publicly available. Access to the UK Biobank data can be requested through a standard protocol (https://www.ukbiobank.ac.uk/register-apply/). The summary statistics of Mendelian randomisation estimates of metabolites and depression can be accessed from https://doi.org/10.1016/j.biopsych.2023.01.027.

The data from NESDA used to support the findings of this study are available upon request from NESDA team, Amsterdam. Information on how to request the study data, including the data sharing policy, can be found at https://www.nesda.nl/nesda-english/.

The data from ALSPAC can be made available on request to the ALSPAC team. Full instructions for applying for data access can be found at http://www.bristol.ac.uk/alspac/researchers/access/. The ALSPAC study website contains details of all the data that are available (http://www.bristol.ac.uk/alspac/researchers/our-data/).

## Code Availability

All codes used for data preparation and analyses are available upon request.

## Acknowledgements

Data from UK Biobank were processed and analysed using RStudio Workbench on the UK Biobank Research Analysis Platform (UKB-RAP), under application #4844. We are extremely grateful to all the UK Biobank, NESDA and ALSPAC participants who took part in this study, to the staff who helped recruit them, and to the wider UK Biobank, NESDA, and ALSPAC teams, including interviewers, computer and laboratory technicians, research scientists, and volunteers. Fig. 1 was created using BioRender.com.

## Author Contributions

ST and XS conceived the study and designed the project. ST developed the computational pipeline, analysed the UK Biobank data, and wrote the original manuscript. DK and ASFK analysed the data in NESDA and ALSPAC, respectively. YM and BWJHP provided consultation and supervised the collaboration with NESDA. MI, AMM, HCW, and XS supervised the project. ETB and GMK provided vital feedback on the initial manuscript. All authors discussed the results, reviewed, and approved the final version.

