## Supplementary 1 for "Metabolome-wide analysis suggests antidepressant exposure counteracts risk of depression in fatty-acid pathways"

### Cohort details

#### UK Biobank (UKB)

Variables: Covariates for the main metabolome-wide association study included age (UKB field: 21003), sex (UKB field: 31), body mass index (BMI, UKB field: 21001), smoking status (UKB field: 20116), assessment centre (UKB field: 54), spectrometer (UKB field: 23650), socioeconomic status (UKB field: 22189), educational qualifications (UKB field: 6138), ethnicity (UKB field: 21000), alcohol intake frequency (UKB field: 1558), alcohol drinker status (UKB field:20117), self-reported depression, and depressive symptoms. Details of all the covariates are provided below.

Socioeconomic status was defined using the Townsend deprivation index scores (UKB field: 22189), which were derived from four indicators of deprivation: unemployment, non-home ownership, non-car ownership, and house overcrowding. Depressive symptoms were measured using PHQ-4 (Patient Health Questionaire-4) questions include: “Frequency of depressed mood in last 2 weeks” (UKB field: 2050), “Frequency of unenthusiasm / disinterest in last 2 weeks” (UKB field: 2060), “Frequency of tenseness / restlessness in last 2 weeks” (UKB field: 2070), “Frequency of tiredness / lethargy in last 2 weeks” (UKB field: 2080). This questionnaire assesses depression-relevant symptoms within a two-week period. The sum of the scores was calculated to indicate the severity of depressive symptoms.

The depression phenotype used in this study was broad depression, which was defined using self-reported help-seeking behaviour for mental health difficulties[1,2]. Case and control status was determined using the touchscreen response to a single item “Seen doctor (GP) for nerves, anxiety, tension or depression” (UKB field: 2090)[1,2]. Cases for broad depression was determined by responding “Yes” to this item or if there was a primary or secondary diagnosis of a depressive mood disorder from linked hospital admission records. Controls were defined if they answered “No”.

In our sensitivity analyses, we additionally adjusted for risk factors included in calculating cardiovascular risk[3], including type 1 and type 2 diabetes, rheumatoid arthritis, kidney failure, kidney nephropathy, glomerulonephritis, migraine, systemic lupus, and erectile dysfunction. These conditions were self-reported by participants at baseline assessment (UKB field: 20002).

#### Netherlands Study of Depression and Anxiety (NESDA)[4]

Cohort description: NESDA is an ongoing cohort study designed to examine the course and consequences of depressive and anxiety disorders. The cohort included 2,981 participants aged 18—65 years, comprising healthy controls, individuals with a prior history of depressive and/or anxiety disorders, and individuals with a current depressive and/or anxiety disorder. Participants were recruited from the community, primary care, and secondary care settings. A detailed description of the NESDA study design and cohort is provided elsewhere[4].

Plasma metabolite profiling: Fasting plasma samples were collected in the morning and stored at -80 °C. Metabolomic profiles were quantified using a proton Nuclear Magnetic Resonance (NMR) platform by Nightingale Health Ltd., Helsinki, Finland. The metabolomic platform measured 51 metabolites with additional sub measures and ratios of lipoproteins (i.e., 98 lipid composition and particle concentration measures of lipoprotein subclasses and 81 lipid and fatty acids ratios)[5]. Of the 125 metabolites selected by Least Absolute Shrinkage and Selection Operator (LASSO) regression in the UKB training cohort, 115 were available in NESDA. Metabolic values were standardised using z-score transformation. Metabolic score was calculated by applying UKB-derived coefficients to the corresponding standardised metabolite levels in NESDA. The score was then used to classify antidepressant exposure using logistic regression, adjusting for age, sex, and shipment batch.

Variables: Sex, age, education level (years), and current smoking status were assessed as part of the clinical interview. BMI (kg/m$^2$) was calculated from weight and height measures. Antidepressant exposure was assessed by inspection of medication containers brought to the assessment, which captured medications used during the preceding month. Recorded medications were subsequently classified according to the Anatomical Therapeutic Chemical classification system to identify antidepressant use. Depression diagnosis was ascertained through the Composite Interview Diagnostic Instrument (CIDI) for major depressive disorder or dysthymia in the past 6 months[6].

In NESDA, we ran three sensitivity analyses when using the metabolic score to classify current antidepressant users from non-current users: (1) restriction to depression cases only; (2) additional adjustment for smoking status, education, and BMI; and (3) restriction to depression cases with additional covariate adjustment.

**Avon Longitudinal Study of Parents and Children (ALSPAC)[7–10]**

Cohort description: Pregnant women resident in Avon, UK with expected dates of delivery between 1st April 1991 and 31st December 1992 were invited to take part in the study. There were 20,248 pregnancies identified as being eligible and the initial number of pregnancies enrolled was 14,541. Of the initial pregnancies, there was a total of 14,676 foetuses, resulting in 14,062 live births and 13,988 children who were alive at 1 year of age. When the oldest children were approximately 7 years of age, an attempt was made to bolster the initial sample with eligible cases who had failed to join the study originally. As a result, when considering variables collected from the age of seven onwards (and potentially abstracted from obstetric notes) there are data available for more than the 14,541 pregnancies mentioned above: The number of new pregnancies not in the initial sample (known as Phase I enrolment) that are currently represented in the released data and reflecting enrolment status at the age of 24 is 906, resulting in an additional 913 children being enrolled (456, 262 and 195 recruited during Phases II, III and IV respectively). The phases of enrolment are described in more detail in the cohort profile paper and its update. The total sample size for analyses using any data collected after the age of seven is therefore 15,447 pregnancies, resulting in 15,658 foetuses. Of these 14,901 children were alive at 1 year of age. From the age of 30, there were an additional 101 new participants recruited from 101 pregnancies during Phase V enrolment. The total sample size for analyses using any data collected after the age of 30 is therefore 15,548 pregnancies, resulting in 15,690 foetuses. Of these 15,002 children were alive at 1 year of age. Study data were collected and managed using REDCap (Research Electronic Data Capture) electronic data capture tools hosted at the University of Bristol. REDCap is a secure, web-based software platform designed to support data capture for research studies[11]. Please note that the ALSPAC study website contains details of all the data that is available through a fully searchable data dictionary and variable search tool: http://www.bristol.ac.uk/alspac/researchers/our-data/. Ethical approval for the study was obtained from the ALSPAC Ethics and Law Committee and the Local Research Ethics Committees. Informed consent for the use of all data collected was obtained from participants following the recommendations of the ALSPAC Ethics and Law Committee at the time. Participants can contact the study team at any time to retrospectively withdraw consent for their data to be used. Study participation is voluntary and during all data collection sweeps, information was provided on the intended use of data. At age 18, study children were sent 'fair processing' materials describing ALSPAC's intended use of their health and administrative records and were given clear means to consent or object via a written form. Data were not extracted for participants who objected, or who were not sent fair processing materials. Ethical approval for the study was obtained from the ALSPAC Law and Ethics committee and local research ethics committees (NHS Haydock REC: 10/H1010/70).

The informed consent obtained from ALSPAC (Avon Longitudinal Study of Parents and Children) participants does not allow the data to be made available through any third party maintained public repository. Supporting data are available from ALSPAC on request under the approved proposal number, B3421. Full instructions for applying for data access can be found here: http://www.bristol.ac.uk/alspac/researchers/access/. The ALSPAC study website contains details of all available data (http://www.bristol.ac.uk/alspac/researchers/our-data/)."

Plasma metabolite profiling: Blood samples were collected at the face-to-face research clinic undertaken at 24 years. Over 220 metabolomic biomarkers (148 metabolites and 77 ratios) were quantified using a high-throughput NMR spectroscopy-based platform (Nightingale Health, Helsinki, Finland). Of the 125 metabolites selected by LASSO regression in the UKB training cohort, 115 were available in ALSPAC. Metabolic values were standardised using z-score transformation. Metabolic was calculated by applying UKB-derived coefficients to the corresponding standardised metabolite levels in ALSPAC. The score was then used to classify antidepressant exposure using logistic regression, adjusting for sex.

Variables: Sex, BMI, and smoking pattern were recorded at the clinic visit at age 24. Age was not included as a covariate as all participants were aged 24 at the time of blood collection. We adopted two definitions of antidepressant case group. First, case group was defined as a "Yes" response to a self-reported questionnaire administered during the blood draw visit, specifically whether the participant was currently taking antidepressants. The control group was defined by a negative response to the same questionnaire items. Additionally, we incorporated a second self-reported questionnaire administered at age 28, which asked participants whether they had been prescribed medication for depression or anxiety within the past five years. In the additional models, case group was defined as a “Yes” response to either of two self-reported items: (1) antidepressant use at age 24, or (2) prescribed medication for depression or anxiety within the past five years, assessed at age 28. The control group in the additional models was defined by negative responses to both self-reported items. Depression phenotype was assessed at age 24 during clinic interview using International Classification of Diseases (ICD)-10 framework[12].

Finally, we conducted three sensitivity analyses in ALSPAC when using the metabolic score to classify cases from controls: (1) restriction to depression cases only; (2) additional adjustment for smoking status and BMI; and (3) restriction to depression cases with additional covariate adjustment.

### Supplementary Methods

##### Basic models

To evaluate collider bias introduced by depression status or depressive symptoms, we conducted the three case/control metWAS using only age, sex, assessment centre, and spectrometer.

##### Sensitivity analyses

In the UKB, we conducted two sensitivity analyses for three comparison groups (current/never, current/past, and current/future): (1) restricting the main models with depression diagnosis, and (2) further adjusting the main models for cardiovascular factors derived from participant’s self-reported medical conditions at baseline assessment (UKB field: 20002). The conditions included as covariates were type 1 diabetes, type 2 diabetes, rheumatoid arthritis, atrial fibrillation, kidney failure, kidney nephropathy, glomerulonephritis, kidney nephropathy, glomerulonephritis, migraine, systemic lupus, and erectile dysfunction.

##### Principal component analysis (PCA)

PCA was performed using the ‘psych’ package (version 2.6.4)[13] to identify dominant patterns among metabolites associated with antidepressant exposure. Metabolites that were significantly associated with antidepressant exposure across all three main metWAS models were included in the PCA. To identify the main contributors to each PC, we examined correlation loadings with absolute values greater than 0.3.

##### G-formula

The parametric g-formula can be used to estimate causal effects of time-varying treatment exposures in the presence of time-varying confounders[15]. As the UKB cohort contained longitudinal data on antidepressant exposure and metabolic profiles measured at baseline assessment and a repeat visit, we applied the parametric g-formula using 'gFormulaMI' package[16] to estimate the effects of time-varying status of antidepressant exposure on metabolic levels.

This analytical approach simulated how each participant's covariates and metabolite levels would change over follow-up under an alternative scenario, also known as regime. In our analyses, we defined the hypothetical exposure regimes of interest as: (1) sustained non-exposure: a scenario in which all participants were not exposed to antidepressant at baseline and repeat assessments, and (2) new exposure: no antidepressant at baseline assessment but exposed to antidepressants at the repeat visit. As such, the estimated effects were derived from the difference in mean metabolite levels between a regime where no one was exposed to antidepressants at every time point and a regime where everyone was exposed to antidepressants at repeat assessment.

Age and BMI at baseline and repeat assessments were included as time-varying covariates, while sex and baseline metabolic level were included as baseline covariates. Metabolite levels at baseline assessment and repeat visit were standardised separately using z-score transformation. Individuals with missing data were excluded. Within 'gFormulaMI', we defined $M$ = 50 imputations, and the number of simulated individuals matched the observed sample size in our data (n = 5,820). Effects and confidence intervals (CI) were derived using pooling rules for synthetic datasets[16].

#### Supplementary Results

##### Basic models

We conducted case-control comparisons between current antidepressant users (cases) and three control groups. All three case-control comparisons (current users versus never users, current users versus past users, and current users versus future users) showed significant differences in the metabolite levels. After adjusting for age, sex, and technical covariates, the basic metWAS models identified 204 (81%) metabolites that showed significant differences between current users and all three control groups after false discovery rate (FDR) correction for multiple testing ($P_{FDR}$ < .05). The highest number of significant associations were found in the current/never comparison (226 significant associations, β = -0.395–0.402, $P_{FDR}$ = < 2.17×10-317 to 0.0380), and the current/future comparison (226, β = -0.261–0.262, $P_{FDR}$ = 1.98×10-111 to 0.0484), then the current/past comparison (221, β = -0.239–0.249, $P_{FDR}$ = 7.49×10-110 to 0.0445). The basic models (Table S5—S7 in Supplement 2) showed largely consistent results, with 92% of metabolites significant in the main models remained significant ($P_{FDR}$ < .05).

#### Sensitivity analyses

In UKB, we first conducted the main models but restricted to participants who had broad depression diagnosis. One-hundred-forty-nine metabolites showed significant differences between current users and all three control groups ($P_{FDR} < .05$). The highest number of significant associations were found in the current/never comparison (180 significant associations, $\beta$ = -0.173–0.179, $P_{FDR}$ ranged from = $0.801\times10^{-44}$ to 0.0497), and the current/past comparison (171, $\beta$ = -0.162–0.159, $P_{FDR}$ ranged from $1.61\times10^{-33}$ to 0.0484), then the current/future comparison (169, $\beta$ = -0.140–0.148, $P_{FDR}$ ranged from $6.25\times10^{-18}$ to 0.0459). After restricting to participants with broad depression diagnosis, 98% of significant biomarkers remained significant.

After adjusting for broad depression, lifestyle-related confounders, and the cardiovascular factors, 154 metabolites showed significant differences between current antidepressant users and all three control groups ($P_{FDR} < .05$). The highest number of significant associations were found in the current/never comparison (189 significant associations, $\beta$ = -0.163–0.180, $P_{FDR}$ ranged from = $0.358\times10^{-53}$ to 0.0477), and the current/future comparison (184, $\beta$ = -0.130–0.150, $P_{FDR}$ ranged from $4.78\times10^{-20}$ to 0.0489), then the the current/past comparison (172, $\beta$ = -0.160–0.164, $P_{FDR}$ ranged from $1.34\times10^{-33}$ to 0.0462). All the significant biomarkers in the three main models (152) retained their significance even after adjusting for cardiovascular factors.

#### PCA

PCA was performed to identify dominant patterns among the metabolites associated with antidepressant exposure. The first 8 principal components (PCs) explained 90% of the total variance cumulatively. Total triglycerides, triglycerides in small VLDL, and triglycerides in small LDL were the major contributors to PC1, in which all showed higher levels in current users of antidepressants than the three control groups in the main metWAS. Cholesteryl esters in very small VLDL, remnant cholesterol (non-HDL, non-LDL -cholesterol), and cholesterol in very small VLDL were the main contributors to PC2. Phospholipids in HDL, phospholipids in HDL, and total lipids in medium HDL were the major contributors to PC3. After excluding metabolites with correlation loadings less than 0.3, the metabolite groups with the largest number of metabolites contributing to the first 8 PCs were relative lipoprotein lipid concentrations (140), lipoprotein subclasses (92), and fatty acids (43).

#### Antidepressant subclasses: Out-of-sample validation

We also trained separate metabolic profile scores for SSRI/SNRI and TCA users using UKB data. In the internal UKB testing sample, the SSRI/SNRI-trained score significantly distinguished current SSRI/SNRI users from all three control groups (never, past, and future users; OR = 1.17–1.25, ROC AUC = 0.55–0.56, PR AUC = 0.05–0.55, $p = 2.66\times10^{-7}$–$2\times10^{-16}$; Figure 7d). Similarly, the TCA-trained score significantly distinguished current TCA users from all three control groups (OR = 1.33–1.45, ROC AUC = 0.58–0.61, PR AUC = $p < 2\times10^{-16}$; Figure 6e).

Both scores also showed significant cross-subclass classification, suggesting limited subclass specificity of the metabolic profiles. The SSRI/SNRI-trained score distinguished current TCA users from never users (OR = 1.30, ROC AUC = 0.56, PR AUC = 0.04, $p = 2.32\times10^{-5}$; Figure 6c). Meanwhile, the TCA-trained score distinguished current SSRI/SNRI users from never users (OR = 1.18, ROC AUC = 0.55, PR AUC = 0.05, p = 0.000274) (Figure 6b).

##### G-formula

Table S27 showed the estimated standardised mean differences in metabolite levels with 95% CI, comparing simulated scenarios in which all participants were not exposed to antidepressants at any time point (sustained non-exposure) versus exposure at repeat visit (new exposure). Nine metabolic biomarkers showed significant differences in mean metabolite levels between the new exposure regime and the sustained non-exposure regime. Total lipids in chylomicrons and extremely large VLDL had the highest estimated mean difference, with new exposure regime scoring 0.202 standard deviations (SD) higher than the sustained non-exposure regime (95% CI = [0.068, 0.336], p = 0.004), followed by triglycerides in chylomicrons and extremely large VLDL (estimated difference = 0.195, 95% CI = [0.073, 0.318], p = 0.002), then phospholipids in chylomicrons and extremely large VLDL (estimated difference = 0.192, 95% CI = [0.042, 0.342], p = 0.0134). Seven out of the nine significant metabolic biomarkers measured extremely large VLDL and its lipid components. The direction of estimates from the g-formula model was consistent with the three main metWAS models (current/never, current/past, current/future) and linear mixed effects models.

#### Supplementary Figures

**Figure S1. Metabolic signatures for current/never, current/past and current/future in main metWAS models. (a) Venn diagram of number of FDR-significant metabolites found for each model and the overlapping significant biomarkers among three models. (b-c) Correlation of effect sizes between the three models. The colour reflects the metabolite group, solid dots indicated FDR–corrected significant in all three models, hollow dots indicated not FDR–corrected significant in all three models.**

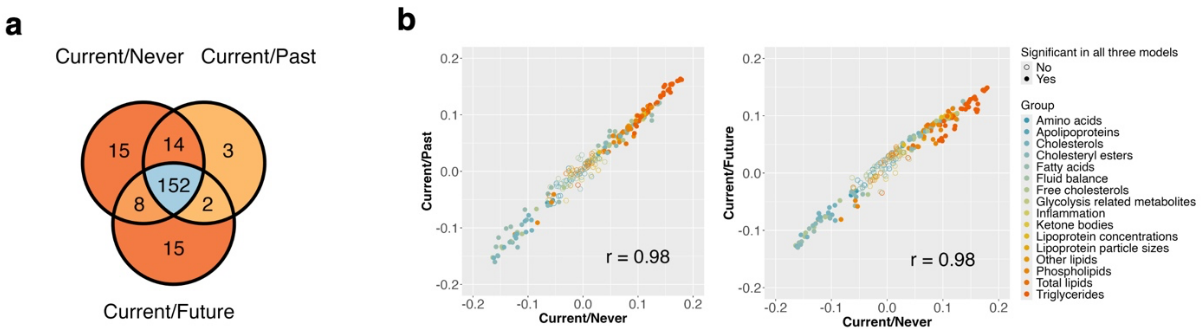

**Figure S2. Forest plots of effect sizes for non-lipoprotein biomarkers from the main current/never comparison model. Hollow dot denotes non-significant associations ($P_{FDR}$ > 0.05).**

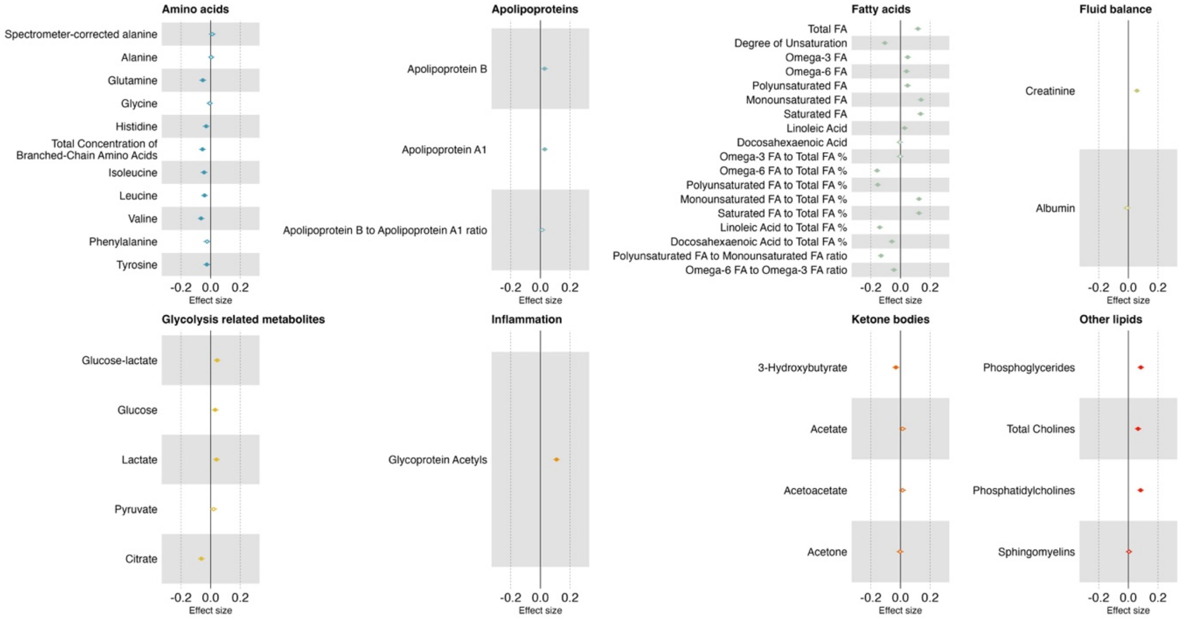

**Figure S3. Comparisons between metabolic signatures of SSRIs/SNRIs and TCAs. (a) Correlation plot of effect sizes from SSRI/SNRIs and TCA in Current/Never exposure. Each dot signifies a metabolic marker, and the colours of the dots indicate whether the metabolites were associated with exposure of both antidepressant subclasses, neither subclasses, SSRIs/SNRIs only, or TCAs only. (b) ROC plot of using SSRI/SNRI and TCA scores to classify Current/Never users of SSRIs/SNRIs. (c) ROC plot of using SSRI/SNRI and TCA scores to classify Current/Never users of TCAs. (d) Forest plot of odds ratios from logistic regression models classifying SSRIs/SNRIs. (e) Forest plot of odds ratios from logistic regression models classifying TCAs.**

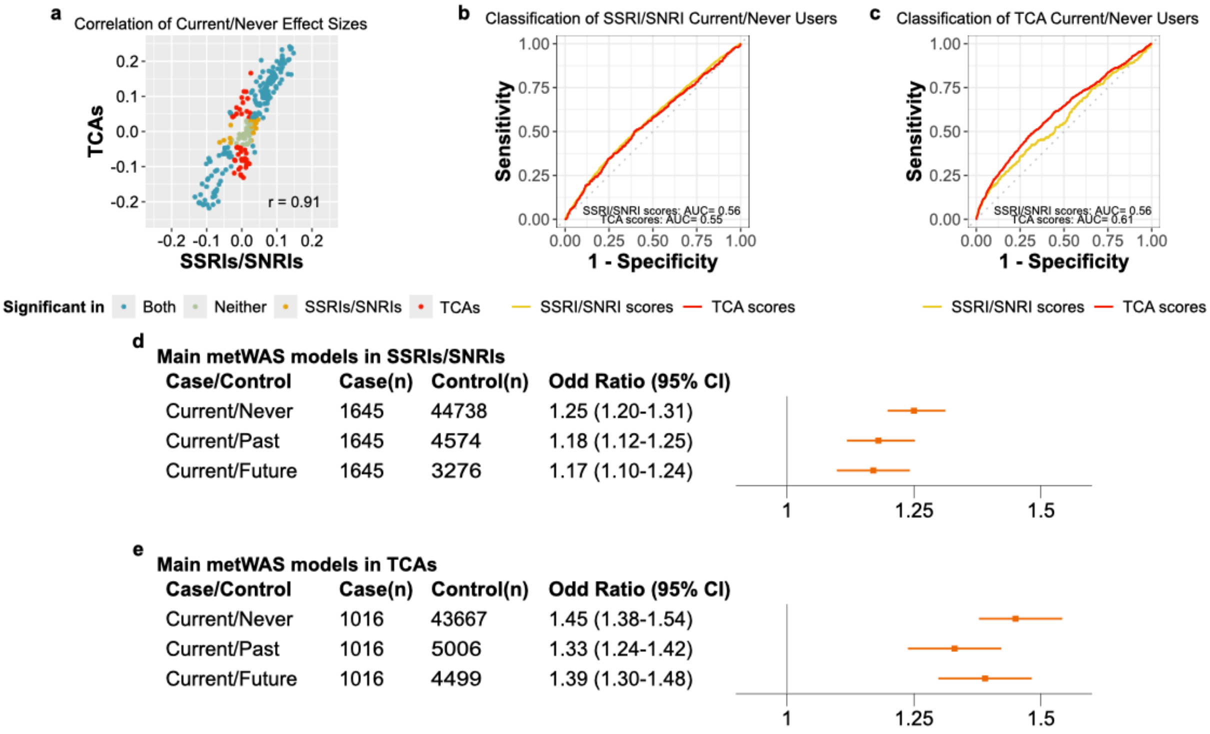

**Figure S4. Heatmaps of effect sizes from the SSRI/SNRI current/never comparison model. NS denotes non-significant ($P_{FDR}$ > .05) associations, and N/A denotes data not available. The 14 lipoprotein subclasses were measured in terms of triglycerides, phospholipids, total cholesterol, cholesteryl esters, and free cholesterol, and relative lipid concentration within each subclass. (a) Biomarkers measuring different lipid components of VLDL, LDL, and HDL. (b) 14 lipoprotein subclasses with their lipid component measured, along with total cholesterol, total cholesteryl esters (esterified cholesterol), total free cholesterol, total phospholipids in lipoprotein particles, and total triglycerides. (c) 14 lipoprotein biomarkers in the relative lipoprotein lipid concentration**

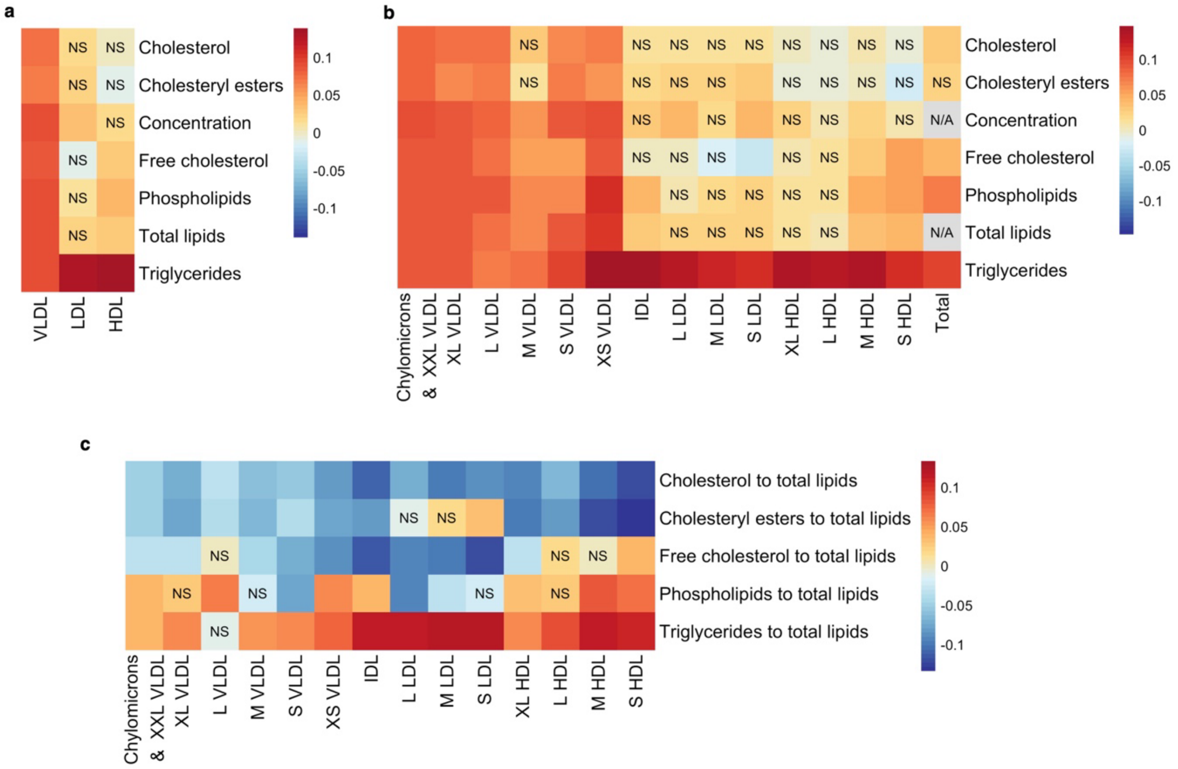

**Figure S5. Forest plots of effect sizes for non-lipoprotein biomarkers from the SSRI/SNRI current/never comparison model. Hollow dot denotes non-significant associations ($P_{FDR}$ > 0.05).**

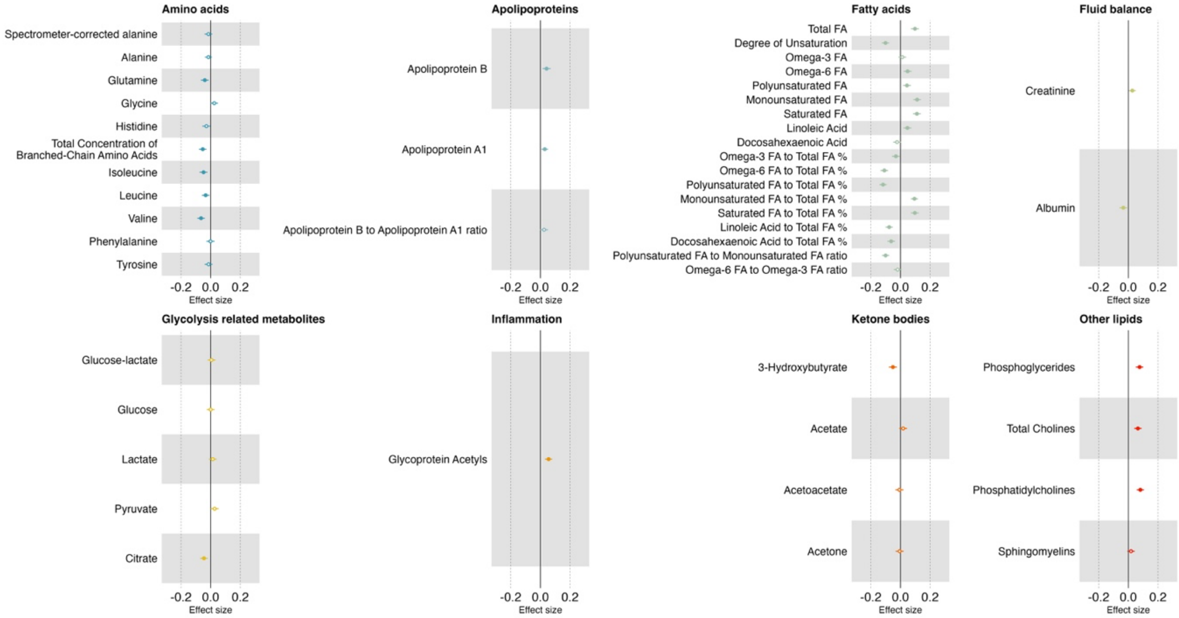

**Figure S6. Heatmaps of effect sizes from the TCA current/never comparison model. NS denotes non-significant ($P_{FDR}$ > .05) associations, and N/A denotes data not available. The 14 lipoprotein subclasses were measured in terms of triglycerides, phospholipids, total cholesterol, cholesteryl esters, and free cholesterol, and relative lipid concentration within each subclass. (a) Biomarkers measuring different lipid components of VLDL, LDL, and HDL. (b) 14 lipoprotein subclasses with their lipid component measured, along with total cholesterol, total cholesteryl esters (esterified cholesterol), total free cholesterol, total phospholipids in lipoprotein particles, and total triglycerides. (c) 14 lipoprotein biomarkers in the relative lipoprotein lipid concentration group**

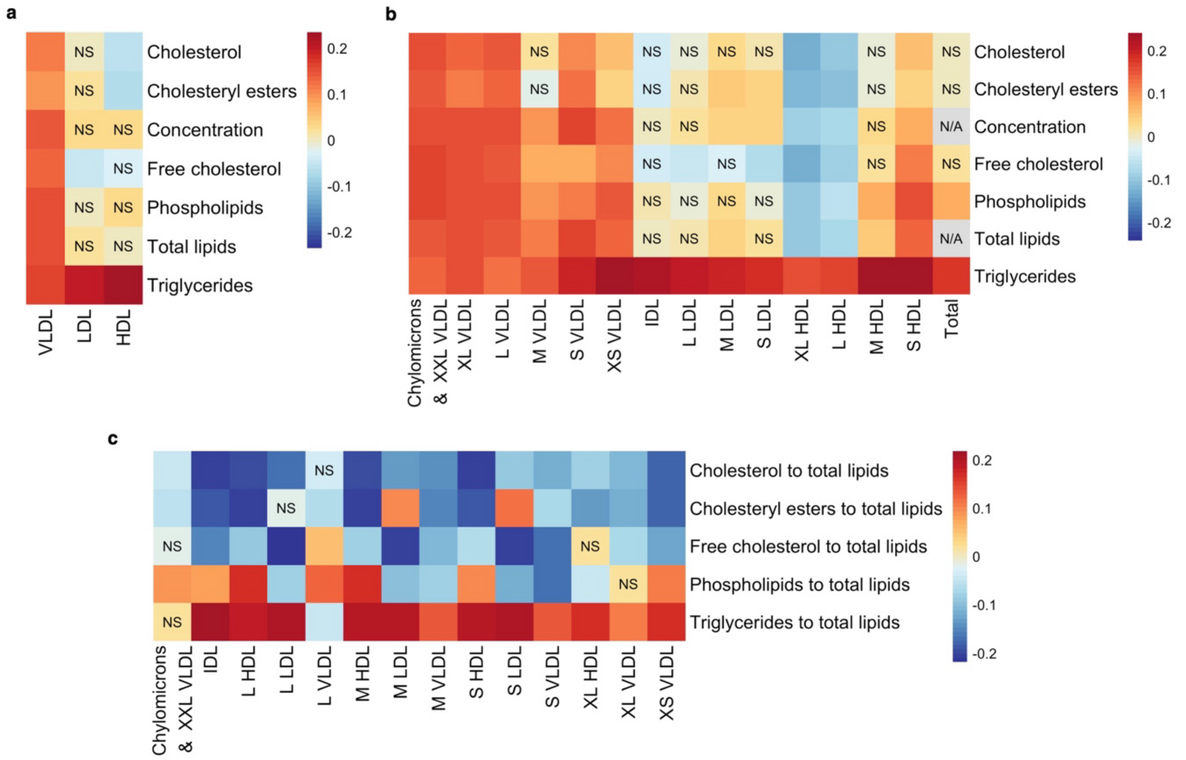

**Figure S7. Forest plots of effect sizes for non-lipoprotein biomarkers from the TCA current/never comparison model.**

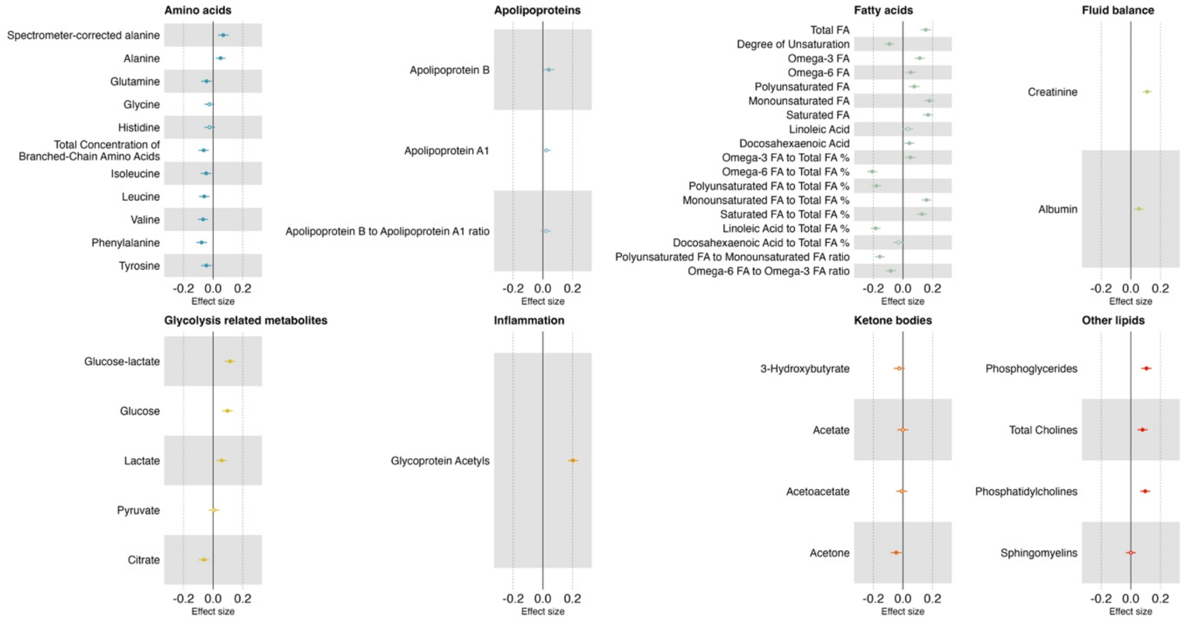

### Supplementary Tables

**Table S1. Comparison of the effect directions between current/never model and MR analyses across metabolite groups. Total number of biomarkers and the percentages of biomarkers showing opposite and same effect directions within each group are presented. Chi-square goodness-of-fit and exact binomial tests were two-sided.**

| **Group** | **Total, n** | **Opposite direction, n (%)** | **Same direction, n (%)** | **$\chi^2$ statistic** | **$\chi^2$ *p*-value** | **Binomial *p*-value** |
|---|---|---|---|---|---|---|
| Triglycerides | 32 | 18.8 | 81.3 | 12.5 | 0.0004 | 0.001 |
| Total lipids | 18 | 77.8 | 22.2 | 5.56 | 0.018 | 0.031 |
| Free cholesterols | 31 | 29.0 | 71.0 | 5.45 | 0.020 | 0.029 |
| Cholesterols | 34 | 32.4 | 67.6 | 4.24 | 0.040 | 0.058 |
| Other lipids | 4 | 100.0 | 0.0 | 4.00 | 0.046 | 0.125 |
| Apolipoproteins | 3 | 100.0 | 0.0 | 3.00 | 0.083 | 0.250 |
| Lipoprotein particle sizes | 3 | 0.0 | 100.0 | 3.00 | 0.083 | 0.250 |
| Cholesteryl esters | 31 | 35.5 | 64.5 | 2.61 | 0.106 | 0.150 |
| Lipoprotein concentrations | 18 | 66.7 | 33.3 | 2.00 | 0.157 | 0.238 |
| Phospholipids | 32 | 40.6 | 59.4 | 1.13 | 0.289 | 0.377 |
| Amino acids | 4 | 75.0 | 25.0 | 1.00 | 0.317 | 0.625 |
| Fatty acids | 15 | 40.0 | 60.0 | 0.60 | 0.439 | 0.607 |

**Table S2. Comparison of the effect directions between SSRI/SNRI current/never model and MR analyses across metabolite groups. Total number of biomarkers and the percentages of biomarkers showing opposite and same effect directions within each group are presented. Chi-square goodness-of-fit and exact binomial tests were two-sided.**

| Group | Total, n | Opposite direction, n (%) | Same direction, n (%) | χ² statistic | χ² *p*-value | Binomial *p*-value |
|---|---|---|---|---|---|---|
| Amino acids | 4 | 75.00 | 25.00 | 1.00 | 0.317 | 0.625 |
| Apolipoproteins | 3 | 100.00 | 0.00 | 3.00 | 0.083 | 0.250 |
| Cholesterols | 34 | 44.12 | 55.88 | 0.47 | 0.493 | 0.608 |
| Cholesteryl esters | 31 | 41.94 | 58.06 | 0.81 | 0.369 | 0.473 |
| Fatty acids | 15 | 40.00 | 60.00 | 0.60 | 0.439 | 0.607 |
| Free cholesterols | 31 | 51.61 | 48.39 | 0.03 | 0.857 | 1.000 |
| Lipoprotein concentrations | 18 | 77.78 | 22.22 | 5.56 | 0.018 | 0.031 |
| Lipoprotein particle sizes | 3 | 66.67 | 33.33 | 0.33 | 0.564 | 1.000 |
| Other lipids | 4 | 100.00 | 0.00 | 4.00 | 0.046 | 0.125 |
| Phospholipids | 32 | 56.25 | 43.75 | 0.50 | 0.480 | 0.597 |
| Total lipids | 18 | 88.89 | 11.11 | 10.89 | 0.001 | 0.001 |
| Triglycerides | 32 | 18.75 | 81.25 | 12.50 | 0.000 | 0.001 |

**Table S3. Comparison of the effect directions between TCA current/never model and MR analyses across metabolite groups. Total number of biomarkers and the percentages of biomarkers showing opposite and same effect directions within each group are presented. Chi-square goodness-of-fit and exact binomial tests were two-sided.**

| Group | Total, n | Opposite direction, n (%) | Same direction, n (%) | χ² statistic | χ² *p*-value | Binomial *p*-value |
|---|---|---|---|---|---|---|
| Amino acids | 4 | 75.00 | 25.00 | 1.00 | 0.317 | 0.625 |
| Apolipoproteins | 3 | 100.00 | 0.00 | 3.00 | 0.083 | 0.250 |
| Cholesterols | 34 | 41.18 | 58.82 | 1.06 | 0.303 | 0.392 |
| Cholesteryl esters | 31 | 35.48 | 64.52 | 2.61 | 0.106 | 0.150 |
| Fatty acids | 15 | 53.33 | 46.67 | 0.07 | 0.796 | 1.000 |
| Free cholesterols | 31 | 22.58 | 77.42 | 9.32 | 0.002 | 0.003 |
| Lipoprotein concentrations | 18 | 61.11 | 38.89 | 0.89 | 0.346 | 0.481 |
| Lipoprotein particle sizes | 3 | 0.00 | 100.00 | 3.00 | 0.083 | 0.250 |
| Other lipids | 4 | 100.00 | 0.00 | 4.00 | 0.046 | 0.125 |
| Phospholipids | 32 | 40.63 | 59.38 | 1.13 | 0.289 | 0.377 |
| Total lipids | 18 | 72.22 | 27.78 | 3.56 | 0.059 | 0.096 |
| Triglycerides | 32 | 18.75 | 81.25 | 12.50 | 0.0004 | 0.001 |
